# Leveraging genomic biobanks to enhance genetic testing outcomes for kidney disease

**DOI:** 10.1101/2025.11.07.25339257

**Authors:** Gretchen M. Urban, Kulsoom Mohammad, Lakshna Sankar, Meei-Hua Lin, Bryn Moore, Mohammad Alkhateeb, Badar Shah, Judith Savige, Peter Harris, Marc S. Williams, Tomohiko Yamamura, Tomoko Horinouchi, Kandai Nozu, Jeffrey H. Miner, Ana Morales, Alexander Chang

## Abstract

Chronic kidney disease (CKD) affects 9.1% globally and is associated with significant morbidity and mortality. While up to 10% of CKD cases yield a genetic diagnosis with hundreds of implicated genes included in some genetic test panels, the results often yield many variants of uncertain significance (VUS). We sought to report genetic testing outcomes in a cohort of 228 patients who received genetic testing for kidney disease phenotypes from 2020-2023 at Geisinger, a regional health system in Pennsylvania. We report on diagnostic yield, management changes, and VUS resolution. A multidisciplinary approach was undertaken to resolve VUS. All cases were reviewed with ≥2 nephrologists and a genetic counselor. VUS with potential clinical relevance, based on known gene-disease associations, were investigated through comprehensive review of genetic databases, consultations with ClinGen variant classification expert panels and genetic testing laboratories, functional assays for Alport syndrome VUS, and case-control analyses using data from a large exome sequencing study (Geisinger MyCode DiscovEHR) linked to electronic health records. Out of 228 patients in this study, 34% had a positive (pathogenic [P] or likely pathogenic [LP]) result, 52% had a VUS result only, and 14% had a negative result. Overall, the mean number of VUS reported was 4.0 (SD 3.1) per patient. After multidisciplinary review of variants, 42 potentially relevant VUS were re-examined by our team. Utilizing MyCode biobank data with available functional analysis we were able to recommend upgrading 10 VUS to LP, which was consistent with genetic laboratories’ decisions to upgrade 5 variants over the course of the study. In addition, our data supported downgrading 10 VUS to likely benign. Genetic testing resulted in direct management changes for 69 (88.5%) positive patients and provided a better understanding of patients’ diagnoses. Genetic testing also had familial implications in 86 (37.7%) of patients. In conclusion, a multidisciplinary approach using biobank data integrating exome sequencing and electronic health records, functional testing, and collaboration with genetic testing laboratories can support the reclassification of clinically significant VUS, potentially maximizing the clinical utility of genetic testing with important diagnostic, prognostic, and management implications.

## Introduction

Kidney diseases can be caused by many different factors, including environmental, lifestyle, and genetic. Due to the heterogenous nature of kidney diseases, identifying monogenic causes can be challenging, particularly as our knowledge of genetic contributors to disease expands and includes milder and, in many cases, overlapping phenotypic presentations. Over 600 genes have been implicated in monogenic kidney diseases.^1,2^ According to the KDIGO’s 2024 Clinical Practice Guideline for the Evaluation and Management of Chronic Kidney Disease, some studies show that >10% of CKD can be attributed to monogenic variants, thus genetic testing should be considered with the standard clinical workup of CKD to establish the cause of CKD.^3^ A National Kidney Foundation Multidisciplinary Working Group of experts recommended genetic testing for a wide variety of clinical scenarios (**Table 1**).

**Table 1.** Indications for Genetic Testing of Kidney Disease.

| Table 1. Indications for Genetic Testing of Kidney Disease |  |
| --- | --- |
| Personal history | • Symptomatic individual with affected first-degree relative<br>• Multi-organ syndromes of unknown etiology<br>• Kidney biopsy findings suggestive of a genetic cause<br>• Atypical clinical presentation where genetic diagnosis may guide therapy<br>• CKD or ESKD of unknown etiology (diagnosed before age 50)<br>• CKD or ESKD of unknown etiology (blood relative considering kidney donation)<br>• CKD or ESKD of unknown etiology (genetic diagnosis may aid in managing extra-renal manifestations)<br>• Atypical cystic kidney or liver disease without a family history |
| Family history | • Familial variant testing for known familial variant<br>• No known familial variant and clinical suspicion exist |
| Evaluation of potential kidney donors | • Donors with a family history of known genetic kidney disease in at least 1 first-degree relative.<br>• Donors with a family history of suspected genetic kidney disease (increased likelihood in case of early onset, extrarenal features, and/or multiple family affected) in at least 1 first-degree relative.<br>• Donors with a family history of kidney disease of unknown etiology in at least 1 first-degree relative or multiple second-degree relatives.<br>• Donors with a family history of known or possible X-linked genetic disease in at least 1 second-degree male relative |

Adapted from the 2024 AJKD article “Advancing Genetic Testing in Kidney Diseases: Report From a National Kidney Foundation Working Group” and AJT article “Genetic Evaluation of living kidney donor candidates”.<sup>42,43</sup>

Following the landmark Groopman et al. (2019) publication, which identified monogenic causes in 9.3% of patients with kidney disease across two cohorts using exome sequencing, genetic testing for kidney disease has increased substantially.^2^ Previously, genetic testing for kidney disease, even clear-cut cases such as Alport syndrome,^4^ was often not performed or only obtained after misdiagnosis and multiple referrals to different institutions. However, the use of genetic testing has surged with the availability of next generation sequencing panels tailored to CKD.^5–8^ A notable tradeoff from this approach is the increased number of variants of uncertain significance (VUS) identified.^8^ A recent study evaluating the clinical utility of genetic testing, using a 382 gene kidney disease panel, reported an average of 7 VUS per patient (range 1-18).^8^

Multidisciplinary efforts are critical in helping resolve VUS, requiring clinicians to work closely with geneticists, genetic counselors, and diagnostic laboratories. The American College of Medical Genetics and Genomics (ACMG) provides criteria for variant classification, and several groups have reported the importance of dedicated VUS follow-up through careful review of literature, in-silico prediction tools, and cosegregation analysis (i.e. measuring how an allele and disease are inherited together in a pedigree).^9–13^ The eMERGE III project published a proof of concept paper that examined how ascertainment of additional data from clinical laboratories, literature, and updated guidelines can be used for variant reclassification; however, this approach only resulted in reclassification of 2% of VUS ^13^. Additional evidence that can aide in reclassification of VUS to pathogenic or benign includes functional testing and case-control studies.^14^

Biobanks with associated medical records offer an opportunity to provide case-control data to help resolve VUS.^12^ Geisinger’s MyCode® Community Health Initiative (MyCode) is a large-scale genomic research initiative within a health system in central and northeast Pennsylvania that integrates genetic data with clinical health records.^15^ In recent years, MyCode data helped establish evidence for protein-truncating variants within the genes *ALG8* and *ALG9* being associated with increased risk of mild cystic kidney disease.^16,17^ Data from biobanks such as the United Kingdom 100,000-genome project, UK Biobank and MyCode have also shown intermediate risk variants contributing significantly to nephrolithiasis with heterozygous *SLC34A3* pathogenic variants associated with 2-3x fold higher risk of nephrolithiasis.^18,19^ Not only do these findings expand our understanding of gene-disease associations, they also provide population-level case-control data that can support the reclassification of VUS in known disease genes.

Clinician engagement is a critical component of the VUS resolution process. Detailed phenotyping, family history collection, and targeted family studies conducted by clinicians can offer case-specific evidence that is not easily available to clinical laboratories. Close collaboration with diagnostic laboratories facilitates sharing these data to perform reanalysis and participate in variant interpretation discussions. These connections, when utilized in conjunction with resources like MyCode, which offers population data relevant to variant interpretation, can create a framework to determine VUS clinical significance.

In this study, we examine genetic kidney disease testing outcomes in a large regional health system from 2020 to 2023, utilizing a multidisciplinary team approach and leveraging Geisinger’s MyCode study and functional testing to try to solve VUS.

## Methods

The study was approved by the Geisinger Institutional Review Board (2022-0312). A total of 228 patients 18 years or older who pursued clinical genetic testing for kidney disorders at Geisinger between 1/1/2020 and 4/1/2023 were included in this study. Geisinger electronic health record (EHR) reviews were conducted by at least 2 nephrologists and 1 genetic counselor to collect information on demographics, indication for genetic testing, clinical history of CKD, clinical features, the specific genetic panel used, genetic testing results, and management outcomes related to genetic testing results.

The genetic test results were categorized into three groups: positive, VUS only, and negative. A positive result was defined as a pathogenic or likely pathogenic variant (P/LP) in an autosomal dominant or X-linked gene or two P/LP variants in an autosomal recessive (AR) gene. VUS-only category was defined as those without a positive result who a VUS result reported. Negative results were categorized as those with no positive or VUS results. Diagnostic yield was calculated for patients with positive results.

As broad-based genetic panel testing often contains more than 380 genes, VUS reporting often includes genes unrelated to the patient’s presenting phenotype. In order to identify patients that harbor VUS in genes consistent with their clinical presentation, additional triage of VUS was conducted by a nephrogenetics expert nephrologist (A.C.) and genetic counselor (G.U.). To maximize the potential number of VUS that may be resolved and support the nephrogenetics community, we prioritized VUS in patients who had VUS-only results as well as those who had a positive diagnostic finding but also harbored a VUS consistent with their clinical phenotype (**Figure 1**). Our multidisciplinary team (medical student, genetic counselors, nephrologists) interrogated multiple sources to obtain population allele frequencies and data about variant classification. Sources included ClinVar, ClinGen, gnomAD,^20–22^ bioinformatic prediction tools (REVEL, alpha missense, spliceAI),^23–25^ the Mayo PKD database for PKD variants,^26^ LOVD for Alport syndrome variants^27^, Alamut, and ClinGen variant classification expert panels for ADPKD and Alport syndrome. Functional testing was performed for Alport syndrome VUS using a minigene assay to detect aberrant splicing by the Nozu lab,^28^ and trimerization reporter NanoLuciferase assays for missense variants by the Miner lab (see Supplemental Methods for full details).^29–31^

**Figure 1.**
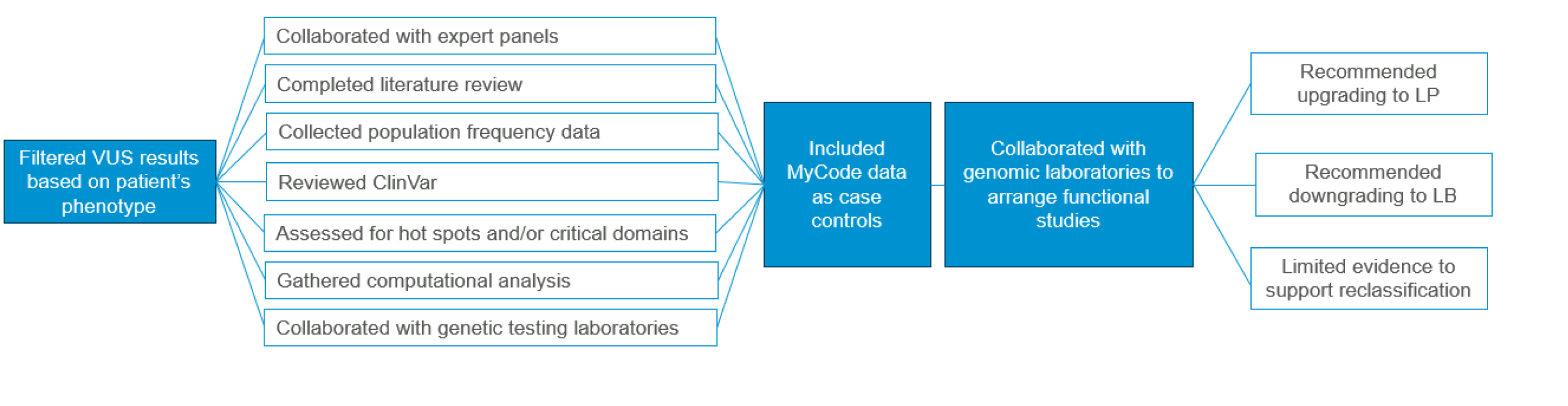
Study Flow.

To assess the frequency of the prioritized VUS in the population, we queried MyCode for individuals harboring each VUS, making sure to exclude the patients from these analyses if they were also in MyCode. Then, for the most common monogenic causes of genetic kidney disease (i.e. *PKD1/PKD2* ADPKD, *COL4A3/4/5* Alport syndrome), we conducted case-control studies comparing VUS heterozygotes vs. non-carriers without rare variants (allele frequency <0.01) in corresponding genes based on our prior work.^32–34^ In addition we examined *SLC34A3* VUS as a risk factor for nephrolithiasis, as we and others have shown that heterozygotes are at mildly increased risk of nephrolithiasis.^18^ For the presence of ADPKD we examined ADPKD ICD codes (see **Supplemental Methods**)^32^. For the presence of Alport syndrome we used dipstick hematuria defined as having trace blood or greater (i.e., trace, 1+, 2+, 3+) on at least half of all urinalyses, excluding urinalyses positive for leukocyte esterase or nitrites.^33,34^ For *SLC34A3,* we used nephrolithiasis phecode GU_585.^35^ We conducted unadjusted chi-square tests or Fisher exact tests (if any cell size ≤5) for each of the VUS, and if significant at a p value threshold of 0.05, we conducted Firth logistic regression analyses adjusting for age, sex, race, ethnicity, initial year in EHR.

Power calculations were based on our prior work using MyCode. ^32–34^ We estimated that we would have >80% power to detect significant differences for case-control analyses if there were at least 5 heterozygotes for *PKD1/PKD2,* 36 for *COL4A3/4/5,* and 178 for *SLC34A3* based on prior analyses using MyCode data (**Supplemental Table 1**). Additional EHR chart reviews were conducted for individuals with VUS of potential clinical significance, focusing on the expected gene-disease relationships and family history.

The number of unrelated probands and number of segregations was collected for each VUS. The number of unrelated probands was determined by identifying individuals with a phenotype consistent with the gene in which the variant was identified. To evaluate for relatedness between cases and evaluate for evidence of family co-segregation, we constructed family pedigrees (1^st^ and 2^nd^ degree relatives) from genetic relatedness data using SimProgeny.^36^ The number of segregations was determined by counting the number of relatives with the same variant and relevant phenotype across families. Efforts were made to avoid double counting probands and families. For example, studies from the same author groups reporting similar cases or pedigrees were flagged for further review or not used in this analysis. We then used these data to reclassify variants if they met criteria per ACMG guidelines.^14^

To determine the clinical outcomes of genetic testing, each case was jointly reviewed by at least 2 nephrology fellows and 2 attending nephrologists from the research team including nephrogenetics experts (A.C., G.U.); final determinations were made through consensus among the reviewers, particularly in instances of initial disagreement. The clinical outcomes of genetic testing were divided into three separate broad categories: changes in management, refining diagnosis and familial implications (**Supplemental Table 2**). To assess changes made to the patient’s medical management, recommendations to avoid extra testing or unnecessary treatment as well as direct personalized treatment and guidance for management of extra renal manifestations were reviewed. The diagnosis was considered refined if genetic testing confirmed a diagnosis, reclassified a diagnosis or aided in the prognosis. Familial implications included the option for cascade testing for family members as well as carrier screening to better understand reproductive risk.

## Results

### Patient demographics and genetic testing information

Out of 228 patients (including 12 potential kidney donors) who received genetic kidney disease testing, 49.1% were female, 84.2% self-reported as White, 9.6% as African American, and 5.7% as Hispanic (**Table 2**). A family history of kidney disease was reported by 67% of patients. Among those with available data, the median (interquartile interval) time from kidney disease diagnosis to genetic testing was 8 (2, 16) years. The majority of initial genetic tests ordered were broad-based panels [n=176 (77.1%)], followed by focused panels [i.e. cystic kidney disease panel or Alport syndrome panel; n=45 (19.7%)], familial variant testing, or *MUC1* testing of the variable-number-of-tandem-repeats (VNTR) region; n=5 (2.2%)], and exome sequencing [n=2(<1%)]. Seventeen (7.5%) underwent multiple genetic tests.

**Table 2.** Patient characteristics.

| Characteristic | N (%) |
| --- | --- |
| Age at kidney diagnosis* | 36.4 (15.3) |
| Age at genetic diagnosis | 46.9 (16.6) |
| Female | 112 (49.1) |
| Self-reported Race or Ethnicity |  |
| White | 192 (84.2) |
| African American | 22 (9.6) |
| Hispanic | 13 (5.7) |
| Pakistani | 1 (0.4) |
| Family history of kidney disease | 153 (67.1) |
| Family history of first degree relative with kidney disease | 133 (58.3) |

\*59 patients missing this data.

### Diagnostic yield of genetic testing and impact of genetic testing on management

Seventy-eight (34%) were found to have a positive result, 32 (14%) were found to have a negative result, and 118 (52%) had a VUS only result (**Figure 2).** The most common indications for genetic testing include cystic kidney disease (n=69, 30%), CKD of unknown etiology (n=40, 18%), nephrolithiasis/nephrocalcinosis (n=24, 11%), and focal segmental glomerulosclerosis (FSGS)/steroid-resistant nephrotic syndrome (SRNS) (n=21, 9%). Diagnostic yield was highest for those tested who had pre-test indications of suspected Alport Syndrome (64%), cystic kidney disease (52%), and tubulointerstitial kidney disease (57%) (**Figure 3**).

**Figure 2.**
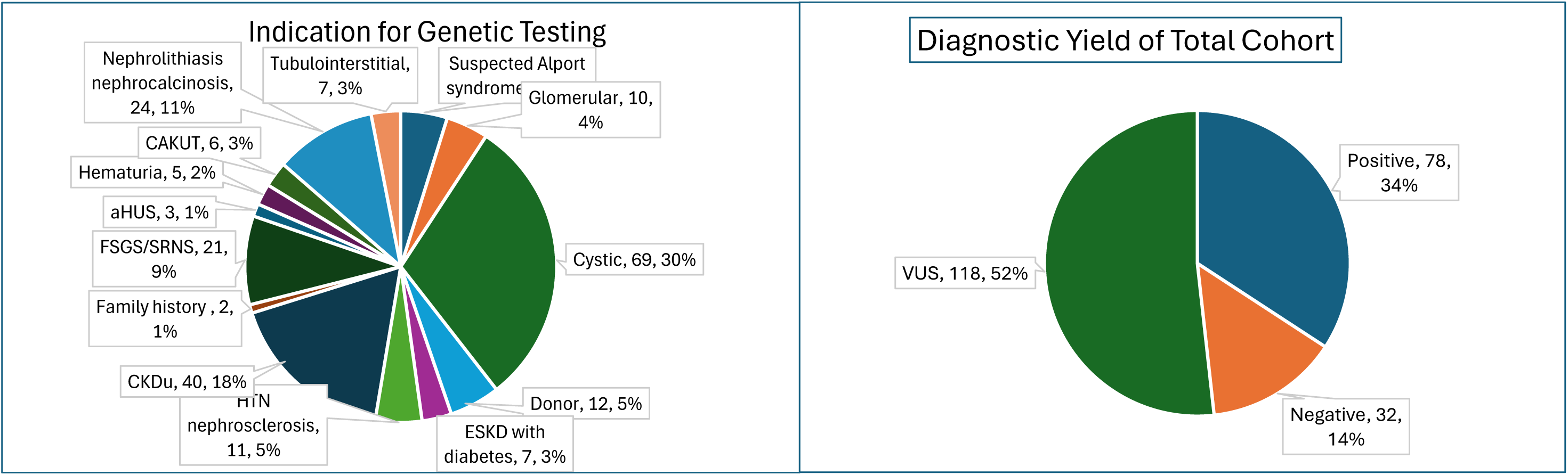
Indication for Genetic Testing and Diagnostic Yield. Abbreviations: HUS, hemolytic uremic syndrome; CAKUT, congenital anomalies of the kidneys and urinary tract; CKDu, chronic kidney disease of unknown etiology; ESKD, end stage kidney disease; FSGS, focal segmental glomerulosclerosis; SRNS, steroid resistant nephrotic syndrome.

**Figure 3.**
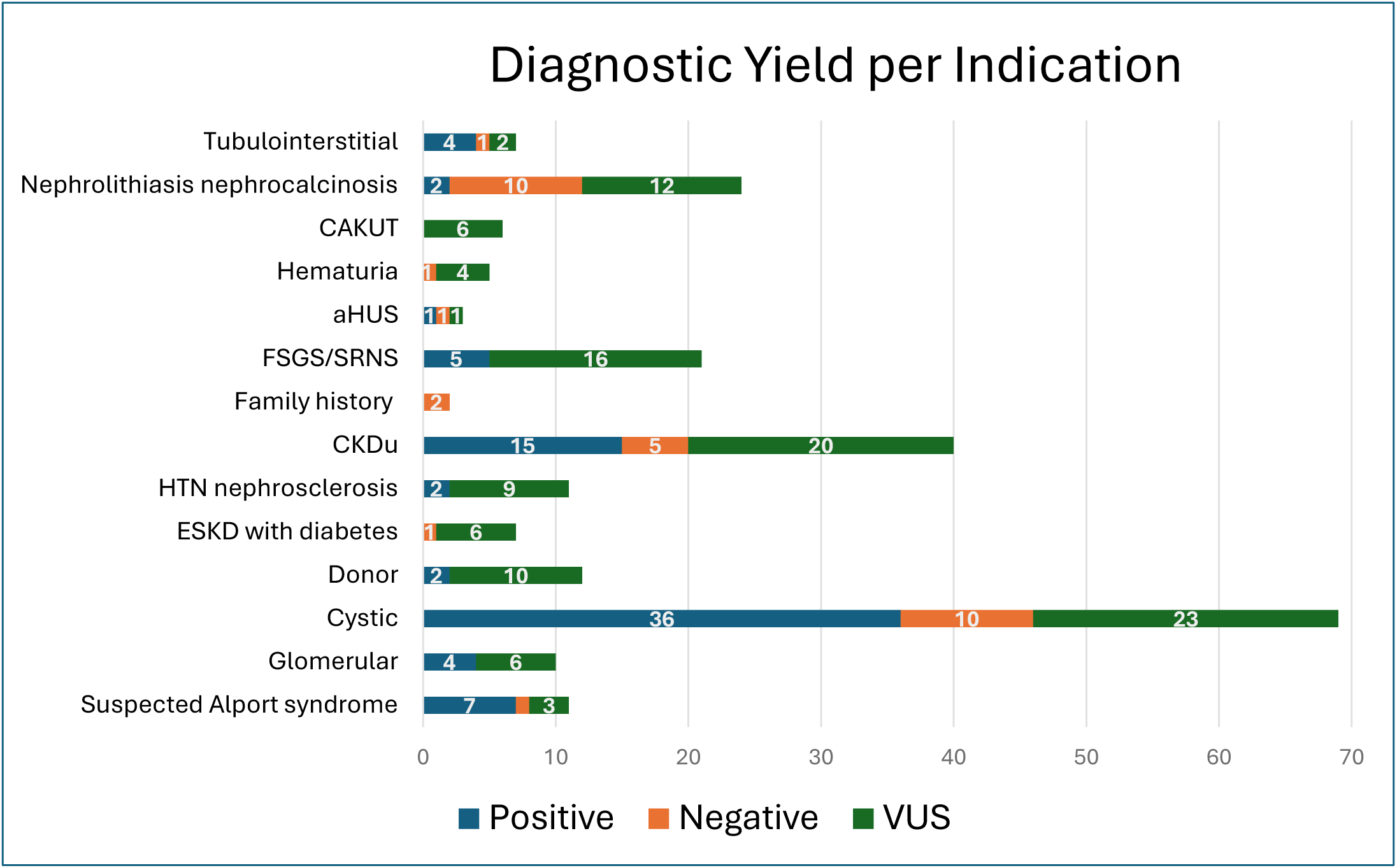
Indication for Genetic Testing and Diagnostic Yield. Abbreviations: HUS, hemolytic uremic syndrome; CAKUT, congenital anomalies of the kidneys and urinary tract; CKDu, chronic kidney disease of unknown etiology; ESKD, end stage kidney disease; FSGS, focal segmental glomerulosclerosis; SRNS, steroid resistant nephrotic syndrome.

When assessing management outcomes, 69 (88.5%) patients had changes to their clinical management. The avoidance of additional testing (e.g. kidney biopsy, unnecessary hematuria workup) was noted for 19 (24.4%) patients, and the avoidance of unnecessary therapy was noted for 24 (30.1%) (**Table 3, Supplemental Table 3)**. Genetic testing results assisted with personalized treatment plans in 37 (47.4%) and provided guidance for management of non-renal manifestations in 55 (70.5%). A more refined diagnosis was provided to patients with a positive result with 78 (100%) receiving a result that confirmed their diagnosis and 35 (45%) who received a result that reclassified their diagnosis.

**Table 3.** Management Outcomes for Positive Results.

| Genetic testing outcomes for patients (N (%)) |  |
| --- | --- |
| Clinical domain | Positives (n=78; 34%) |
| Change in clinical management: | 69 (88.5%) |
| • Avoidance of additional testing | • 19 (24.4%) |
| • Avoidance of unnecessary therapy | • 24 (30.1%) |
| • Personalized treatment plan | • 37 (47.4%) |
| • Management of extrarenal concerns | • 55 (70.5%) |
| Refine diagnosis: | 78 (100%) |
| • Confirmed diagnosis | • 43 (55%) |
| • Reclassified diagnosis | • 35 (45%) |
| • Prognostic clarification | • 45 (57.7%) |

Further, genetic testing led to a better understanding of the patient’s disease prognosis in 45 (57.7%) patients who received a positive result. Each positive result had implications on familial risk by identifying family members who could be affected. Of the total cohort, genetic testing identified carrier status in 86 (37.7%) of patients, which had potential to help relatives better understand their reproductive risk.

### Follow-up evaluation of VUS

After multidisciplinary review (AC and GU) of patients with VUS, there were 42 VUS in genes consistent with the patient’s clinical phenotype that were evaluated further (**Tables 4-6, Supplemental Tables 4-5**). Of these VUS, our review supported upgrading 10 VUS to LP. During the time of the study, 5 of these were upgraded by the genetic testing laboratory, including 3 based on previously published Geisinger MyCode data.^12,32^ Further, we were able to generate evidence to recommend the reclassification of an additional 10 VUS to likely benign (LB).

**Table 4.**
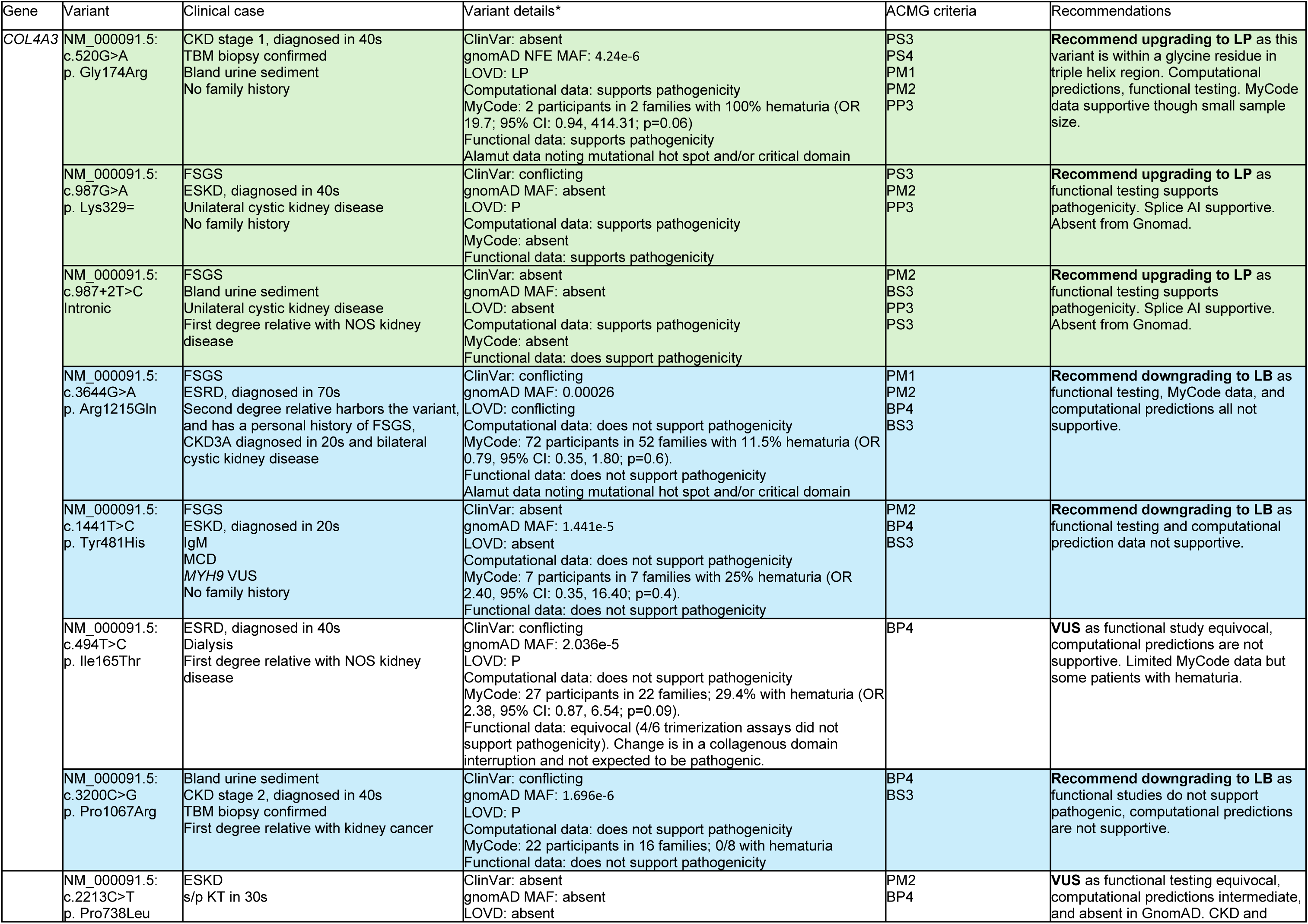
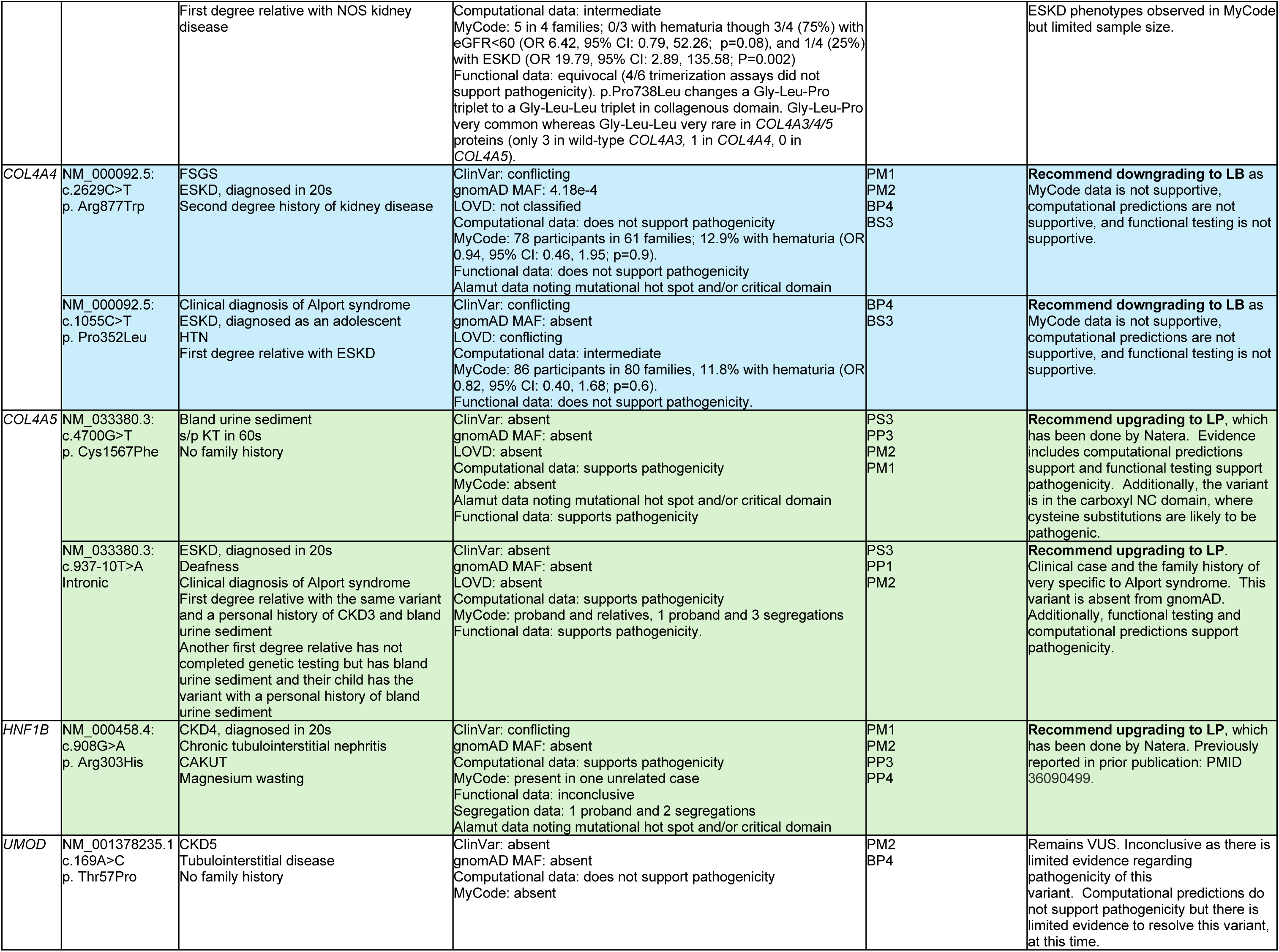
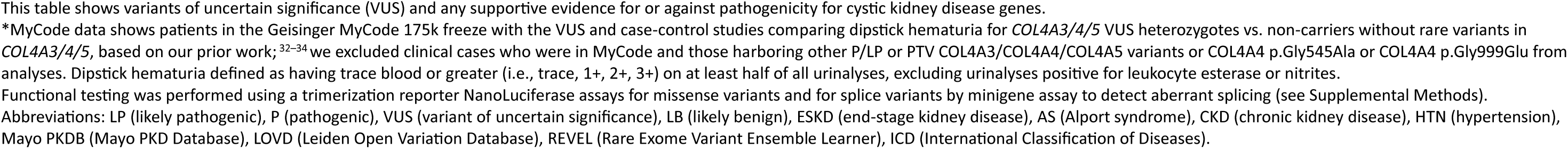
Variants in *COL4A3/4/5, UMOD, and HNF1B* Investigated.

| Gene | Variant | Clinical case | Variant details* | ACMG criteria | Recommendations |
| --- | --- | --- | --- | --- | --- |
| COL4A3 | NM_000091.5:<br>c.520G>A<br>p. Gly174Arg | CKD stage 1, diagnosed in 40s<br>TBM biopsy confirmed<br>Bland urine sediment<br>No family history | ClinVar: absent<br>gnomAD NFE MAF: 4.24e-6<br>LOVD: LP<br>Computational data: supports pathogenicity<br>MyCode: 2 participants in 2 families with 100% hematuria (OR 19.7; 95% CI: 0.94, 414.31; p=0.06)<br>Functional data: supports pathogenicity<br>Alamut data noting mutational hot spot and/or critical domain | PS3<br>PS4<br>PM1<br>PM2<br>PP3 | **Recommend upgrading to LP** as this variant is within a glycine residue in triple helix region. Computational predictions, functional testing. MyCode data supportive though small sample size. |
|  | NM_000091.5:<br>c.987G>A<br>p. Lys329= | FSGS<br>ESKD, diagnosed in 40s<br>Unilateral cystic kidney disease<br>No family history | ClinVar: conflicting<br>gnomAD MAF: absent<br>LOVD: P<br>Computational data: supports pathogenicity<br>MyCode: absent<br>Functional data: supports pathogenicity | PS3<br>PM2<br>PP3 | **Recommend upgrading to LP** as functional testing supports pathogenicity. Splice AI supportive. Absent from Gnomad. |
|  | NM_000091.5:<br>c.987+2T>C<br>Intronic | FSGS<br>Bland urine sediment<br>Unilateral cystic kidney disease<br>First degree relative with NOS kidney disease | ClinVar: absent<br>gnomAD MAF: absent<br>LOVD: absent<br>Computational data: supports pathogenicity<br>MyCode: absent<br>Functional data: does support pathogenicity | PM2<br>BS3<br>PP3<br>PS3 | **Recommend upgrading to LP** as functional testing supports pathogenicity. Splice AI supportive. Absent from Gnomad. |
|  | NM_000091.5:<br>c.3644G>A<br>p. Arg1215Gln | FSGS<br>ESRD, diagnosed in 70s<br>Second degree relative harbors the variant, and has a personal history of FSGS, CKD3A diagnosed in 20s and bilateral cystic kidney disease | ClinVar: conflicting<br>gnomAD MAF: 0.00026<br>LOVD: conflicting<br>Computational data: does not support pathogenicity<br>MyCode: 72 participants in 52 families with 11.5% hematuria (OR 0.79, 95% CI: 0.35, 1.80; p=0.6).<br>Functional data: does not support pathogenicity<br>Alamut data noting mutational hot spot and/or critical domain | PM1<br>PM2<br>BP4<br>BS3 | **Recommend downgrading to LB** as functional testing, MyCode data, and computational predictions all not supportive. |
|  | NM_000091.5:<br>c.1441T>C<br>p. Tyr481His | FSGS<br>ESKD, diagnosed in 20s<br>IgM<br>MCD<br>MYH9 VUS<br>No family history | ClinVar: absent<br>gnomAD MAF: 1.441e-5<br>LOVD: absent<br>Computational data: does not support pathogenicity<br>MyCode: 7 participants in 7 families with 25% hematuria (OR 2.40, 95% CI: 0.35, 16.40; p=0.4).<br>Functional data: does not support pathogenicity | PM2<br>BP4<br>BS3 | **Recommend downgrading to LB** as functional testing and computational prediction data not supportive. |
|  | NM_000091.5:<br>c.494T>C<br>p. Ile165Thr | ESRD, diagnosed in 40s<br>Dialysis<br>First degree relative with NOS kidney disease | ClinVar: conflicting<br>gnomAD MAF: 2.036e-5<br>LOVD: P<br>Computational data: does not support pathogenicity<br>MyCode: 27 participants in 22 families; 29.4% with hematuria (OR 2.38, 95% CI: 0.87, 6.54; p=0.09).<br>Functional data: equivocal (4/6 trimerization assays did not support pathogenicity). Change is in a collagenous domain interruption and not expected to be pathogenic. | BP4 | **VUS** as functional study equivocal, computational predictions are not supportive. Limited MyCode data but some patients with hematuria. |
|  | NM_000091.5:<br>c.3200C>G<br>p. Pro1067Arg | Bland urine sediment<br>CKD stage 2, diagnosed in 40s<br>TBM biopsy confirmed<br>First degree relative with kidney cancer | ClinVar: conflicting<br>gnomAD MAF: 1.696e-6<br>LOVD: P<br>Computational data: does not support pathogenicity<br>MyCode: 22 participants in 16 families; 0/8 with hematuria<br>Functional data: does not support pathogenicity | BP4<br>BS3 | **Recommend downgrading to LB** as functional studies do not support pathogenic, computational predictions are not supportive. |
|  | NM_000091.5:<br>c.2213C>T<br>p. Pro738Leu | ESKD<br>s/p KT in 30s | ClinVar: absent<br>gnomAD MAF: absent<br>LOVD: absent | PM2<br>BP4 | **VUS** as functional testing equivocal, computational predictions intermediate, and absent in GnomAD. CKD and |
|  |  | First degree relative with NOS kidney disease | Computational data: intermediate<br>MyCode: 5 in 4 families; 0/3 with hematuria though 3/4 (75%) with eGFR<60 (OR 6.42, 95% CI: 0.79, 52.26; p=0.08), and 1/4 (25%) with ESKD (OR 19.79, 95% CI: 2.89, 135.58; P=0.002)<br>Functional data: equivocal (4/6 trimerization assays did not support pathogenicity). p.Pro738Leu changes a Gly-Leu-Pro triplet to a Gly-Leu-Leu triplet in collagenous domain. Gly-Leu-Pro very common whereas Gly-Leu-Leu very rare in COL4A3/4/5 proteins (only 3 in wild-type COL4A3, 1 in COL4A4, 0 in COL4A5). |  | ESKD phenotypes observed in MyCode but limited sample size. |
| COL4A4 | NM_000092.5:<br>c.2629C>T<br>p. Arg877Trp | FSGS<br>ESKD, diagnosed in 20s<br>Second degree history of kidney disease | ClinVar: conflicting<br>gnomAD MAF: 4.18e-4<br>LOVD: not classified<br>Computational data: does not support pathogenicity<br>MyCode: 78 participants in 61 families; 12.9% with hematuria (OR 0.94, 95% CI: 0.46, 1.95; p=0.9).<br>Functional data: does not support pathogenicity<br>Alamut data noting mutational hot spot and/or critical domain | PM1<br>PM2<br>BP4<br>BS3 | **Recommend downgrading to LB** as MyCode data is not supportive, computational predictions are not supportive, and functional testing is not supportive. |
|  | NM_000092.5:<br>c.1055C>T<br>p. Pro352Leu | Clinical diagnosis of Alport syndrome<br>ESKD, diagnosed as an adolescent<br>HTN<br>First degree relative with ESKD | ClinVar: conflicting<br>gnomAD MAF: absent<br>LOVD: conflicting<br>Computational data: intermediate<br>MyCode: 86 participants in 80 families, 11.8% with hematuria (OR 0.82, 95% CI: 0.40, 1.68; p=0.6).<br>Functional data: does not support pathogenicity. | BP4<br>BS3 | **Recommend downgrading to LB** as MyCode data is not supportive, computational predictions are not supportive, and functional testing is not supportive. |
| COL4A5 | NM_033380.3:<br>c.4700G>T<br>p. Cys1567Phe | Bland urine sediment<br>s/p KT in 60s<br>No family history | ClinVar: absent<br>gnomAD MAF: absent<br>LOVD: absent<br>Computational data: supports pathogenicity<br>MyCode: absent<br>Alamut data noting mutational hot spot and/or critical domain<br>Functional data: supports pathogenicity | PS3<br>PP3<br>PM2<br>PM1 | **Recommend upgrading to LP**, which has been done by Natera. Evidence includes computational predictions support and functional testing support pathogenicity. Additionally, the variant is in the carboxyl NC domain, where cysteine substitutions are likely to be pathogenic. |
|  | NM_033380.3:<br>c.937-10T>A<br>Intronic | ESKD, diagnosed in 20s<br>Deafness<br>Clinical diagnosis of Alport syndrome<br>First degree relative with the same variant and a personal history of CKD3 and bland urine sediment<br>Another first degree relative has not completed genetic testing but has bland urine sediment and their child has the variant with a personal history of bland urine sediment | ClinVar: absent<br>gnomAD MAF: absent<br>LOVD: absent<br>Computational data: supports pathogenicity<br>MyCode: proband and relatives, 1 proband and 3 segregations<br>Functional data: supports pathogenicity. | PS3<br>PP1<br>PM2 | **Recommend upgrading to LP**. Clinical case and the family history of very specific to Alport syndrome. This variant is absent from gnomAD. Additionally, functional testing and computational predictions support pathogenicity. |
| HNF1B | NM_000458.4:<br>c.908G>A<br>p. Arg303His | CKD4, diagnosed in 20s<br>Chronic tubulointerstitial nephritis<br>CAKUT<br>Magnesium wasting | ClinVar: conflicting<br>gnomAD MAF: absent<br>Computational data: supports pathogenicity<br>MyCode: present in one unrelated case<br>Functional data: inconclusive<br>Segregation data: 1 proband and 2 segregations<br>Alamut data noting mutational hot spot and/or critical domain | PM1<br>PM2<br>PP3<br>PP4 | **Recommend upgrading to LP**, which has been done by Natera. Previously reported in prior publication: PMID 36090499. |
| UMOD | NM_001378235.1<br>c.169A>C<br>p. Thr57Pro | CKD5<br>Tubulointerstitial disease<br>No family history | ClinVar: absent<br>gnomAD MAF: absent<br>Computational data: does not support pathogenicity<br>MyCode: absent | PM2<br>BP4 | Remains VUS. Inconclusive as there is limited evidence regarding pathogenicity of this variant. Computational predictions do not support pathogenicity but there is limited evidence to resolve this variant, at this time. |

This table shows variants of uncertain significance (VUS) and any supportive evidence for or against pathogenicity for cystic kidney disease genes.
\*MyCode data shows patients in the Geisinger MyCode 175k freeze with the VUS and case-control studies comparing dipstick hematuria for *COL4A3/4/5* VUS heterozygotes vs. non-carriers without rare variants in *COL4A3/4/5*, based on our prior work;<sup>32-34</sup> we excluded clinical cases who were in MyCode and those harboring other P/LP or PTV *COL4A3/COL4A4/COL4A5* variants or *COL4A4* p.Gly545Ala or *COL4A4* p.Gly999Glu from analyses. Dipstick hematuria defined as having trace blood or greater (i.e., trace, 1+, 2+, 3+) on at least half of all urinalyses, excluding urinalyses positive for leukocyte esterase or nitrites.
Functional testing was performed using a trimerization reporter NanoLuciferase assays for missense variants and for splice variants by minigene assay to detect aberrant splicing (see Supplemental Methods).
Abbreviations: LP (likely pathogenic), P (pathogenic), VUS (variant of uncertain significance), LB (likely benign), ESKD (end-stage kidney disease), AS (Alport syndrome), CKD (chronic kidney disease), HTN (hypertension), Mayo PKDB (Mayo PKD Database), LOVD (Leiden Open Variation Database), REVEL (Rare Exome Variant Ensemble Learner), ICD (International Classification of Diseases).

Twelve of the VUS investigated were within the *COL4A3/4/5* genes. Of these, we had sufficient MyCode biobank case-control and functional testing data (<75% of wild-type in intracellular and/or extracellular trimer levels using a Trimerization reporter NanoLuciferase assay) to support reclassification of 5 VUS to LP and 5 to LB. For example, a *COL4A3* c.520G>A (p.Gly174Arg) VUS detected in a 40–45-year-old patient with thin basement membrane disease, severe proteinuria, and hematuria had multiple lines of evidence: variant location affecting a glycine residue in triple helix region, supportive computational predictions, case-control biobank data, and functional testing showing reduced secretion (**Table 4**). Another proband with ESKD diagnosed in their 20s and bilateral sensorineural deafness was identified with a *COL4A5* c.937-10T>A intronic variant. The same variant was identified in multiple first degree relatives of the proband. Mini-gene assay demonstrated this intronic variant resulted in abnormal splicing (**Supplemental Figure**). Two variants (*COL4A3* p.Ile165Thr, *COL4A3* p.Pro738Leu) had equivocal functional testing findings with each variant having 4/6 trimerization assays favoring non-pathogenicity. *COL4A3* p.Ile165Thr variant causes a collagenous domain interruption, which is not expected to be pathogenic; limited MyCode data were available for only 27 participants with 29.4% (OR 2.38, 95% CI: 0.87, 6.54; p=0.09) having dipstick hematuria. *COL4A3* p.Pro738Leu is absent in large population databases and results in a change in a Gly-Leu-Pro triplet to Gly-Leu-Leu triplet in the collagenous domain. Whereas Gly-Leu-Pro is very common in *COL4A3/4/5* proteins, Gly-Leu-Leu is exceedingly rare (only 3 in wild-type *COL4A3,* 1 in *COL4A4,* 0 in *COL4A5*). None of the *COL4A3* p.Pro738Leu heterozygotes had hematuria but 3/4 had eGFR<60 ml/min/1.73m^2^ (OR 6.42, 95% CI: 0.79, 52.26; p=0.08), and 1/4 had ESKD (OR 19.79, 95% CI: 2.89, 135.58; p=0.002) (**Table 4**). While the trimerization assay we used is well-validated for demonstrating pathogenicity, ^29–31^ we cannot rule out the possibility of missing some aspect that is functionally important in vivo.

Further, our data support reclassifying 5 *COL43/4/5* VUS as LB, bolstered by case-control data and functional testing. In our MyCode research cohort, individuals with the *COL4A4* p.Arg877Trp (n=78), *COL4A4* p.Pro352Leu (n=88), and *COL4A3* p.Arg1215Gln (n=72) variants were not at increased risk of dipstick hematuria (all p values >0.05 compared to non-carrier controls), along with consistent negative functional testing (**Table 4**). Similarly, there were 4 *PKD1* and 1 *PKD2* VUS, for which our data support reclassifying as LB with large numbers of individuals without ADPKD ICD codes in MyCode (*PKD1* p.Phe2493Leu [0/15 with ADPKD], *PKD1* p.Ser4288Gly [0/65 with ADPKD], *PKD1* p.Arg2765Cys [2/2533 with ADPKD; p=0.6], *PKD1* p.Arg1190Cys [0/11 with ADPKD]; PKD2 p.Thr301Ala [0/12 with ADPKD] (**Table 5)**.

**Table 5.** Variants in Cystic Kidney Disease genes (*PKD1, PKD2, IFT140, TSC1)* Investigated.

| Autosomal Dominant Polycystic Kidney Disease (ADPKD) |  |  |  |  |  |
| --- | --- | --- | --- | --- | --- |
| Gene | Variant | Clinical case | Variant details* | ACMG criteria | Recommendations |
| PKD1 | NM_001009944.3: c.6101G>T<br>p. Gly2034Val | CKD2, diagnosed in 40s<br>Bilateral cystic kidney disease<br>Possible R cerebellar artery aneurysm<br>First degree relative with PKD | ClinVar: VUS<br>Mayo PKDB: absent<br>gnomAD MAF: absent<br>Computational data: support pathogenicity<br>MyCode: 1/3 (33%) had ADPKD ICD code. Each participant had ADPKD and family segregation upon chart review.<br>Alamut data: mutational hot spot and/or critical domain<br>Publication (PMID 36573973): showing evidence of segregation and atypical presentation of ADPKD | PM2<br>PP3<br>PP1<br>PM1 | **Recommend upgrading to LP**, which was done by Natera while this study was being conducted. |
|  | NM_001009944.3: c.1543G>A<br>p. Gly515Arg | CKD1, diagnosed in 30s<br>Cystic kidney disease<br>No family history | ClinVar: conflicting<br>gnomAD MAF: absent<br>Mayo PKDB: LP<br>Computational data: intermediate<br>MyCode: absent<br>Alamut data noting mutational hot spot and/or critical domain | PM1<br>PM2<br>PP3 | **Recommend upgrading to LP**. GeneDx upgraded this variant during the study time due to a patient and his parent having the variant as well as 2 other affected patients in GeneDx database. |
|  | NM_001009944.3: c.7531G>C<br>p. Ala2511Pro | ESRD, diagnosed in 30s<br>Cystic kidney disease<br>Liver cysts<br>3 first degree relatives with cystic kidney, 2 relatives positive for the same *PKD1* variant. | ClinVar: absent<br>gnomAD MAF: absent<br>Mayo PKDB: VUS<br>Computational data: intermediate<br>MyCode: 1/1 (related to clinical case) has an ADPKD ICD code<br>Alamut data noting mutational hot spot and/or critical domain | PM1<br>PM2<br>BP4<br>PP4 | **Natera recently upgraded variant to LP but we recommend downgrading it to VUS due to the possibility that the MyCode participant was double counted.** Therefore, segregation data is inconclusive.<br>Previously reported: (PMID 36573973) |
|  | NM_001009944.3: c.7479C>G<br>p. Phe2493Leu | CKD4, diagnosed in 60s<br>Cystic kidney disease<br>*COL4A1* LP<br>First degree relative with NOS kidney disease, early stroke, muscle cramps, and nephrolithiasis | ClinVar: VUS<br>Mayo PKDB: absent<br>gnomAD MAF: low<br>Computational data: support pathogenicity<br>MyCode: 0/15 participants with ADPKD ICD code<br>Alamut data: mutational hot spot and/or critical domain | PP3<br>BP5<br>BS2<br>PM1<br>PM2 | **Recommend downgrading to LB** given that MyCode data is not supportive of pathogenic and our patient's phenotype overlaps with *COL4A1*-related concerns. However, computational evidence supports a deleterious effect, and the variant is within the PKD/REJ-like protein domain. |
|  | NM_001009944.3: c.12862A>G<br>p. Ser4288Gly | CKD2, diagnosed in 30s<br>Cystic kidney disease<br>*ALG8* P/LP<br>No family history | ClinVar: VUS<br>gnomAD MAF: low<br>Mayo PKDB: LB<br>Computational data: does not support pathogenicity<br>MyCode: 0/65 participants with ADPKD ICD codes | PM2<br>BP4<br>BP5 | **Recommend downgrading to LB** as MyCode data and computational prediction data are not supportive of pathogenicity. Additionally, the patient's phenotype is consistent with *ALG8*-related concerns. |
|  | NM_001009944.3: c.8293C>T<br>p. Arg2765Cys | ESRD, diagnosed in 20s<br>Cystic kidney disease<br>*HNF1B* P<br>First degree relative with history of cystic kidney disease | ClinVar: conflicting data<br>gnomAD MAF: absent<br>Mayo PKDB: VUS<br>Computational data: intermediate<br>MyCode: 2/2533 (0.08%) with ADPKD ICD code (OR 1.45, 95% CI: 0.38, 5.59; p=0.6)<br>Alamut data noting mutational hot spot and/or critical domain | PM1<br>PM2<br>BP4<br>BP5 | **Recommend downgrading to LB**. MyCode data is unsupportive and more common than most *PKD1* variants. Computational prediction data is not supportive. Proband's case can be explained by *HNF1B* pathogenic variant. Could be a modifier variant given the clinical ADPKD history. |
|  |  |  | Publication (PMID 19165178): variant seen with second PKD1 variant in symptomatic individual and predicted to be LP based on coding effect. |  |  |
|  | NM_001009944.3:<br>c.3568C>T<br>p. Arg1190Cys | Cystic kidney disease<br>*PKD2* P<br>No family history | ClinVar: conflicting<br>gnomAD MAF: low<br>Mayo PKDB: absent<br>Computational data: does not support pathogenicity<br>MyCode: 0/11 with ADPKD ICD code.<br>Alamut data noting mutational hot spot and/or critical domain | PM1<br>PM2<br>BP4<br>BP5 | **Recommend downgrading to LB** as MyCode data not supportive and computational prediction data not supportive. Proband's case explained by *PKD2* pathogenic variant. |
|  | NM_001009944.3:<br>c.7622 C>T<br>p. Pro2541Leu | CKD3B, diagnosed in 80s<br>Cystic kidney disease<br>No family history | ClinVar: VUS<br>gnomAD MAF: absent<br>Mayo PKDB: absent<br>Computational data: intermediate<br>MyCode: 0/65 with ADPKD ICD codes; 4/65 with cystic kidney/liver ICD code (OR 2.41, 95% CI: 0.91, 6.38; p=0.08)<br>Alamut data noting mutational hot spot and/or critical domain | PM1<br>PM2 | Remains VUS. Inconclusive as MyCode data without any ADPKD diagnoses but computational prediction data intermediate. |
|  | NM_001009944.3:<br>c.9376A>G<br>p. Thr3126Ala | Fetal CAKUT detected on US<br>Proband and fetus have the variant<br>First and second degree relative with cystic kidney disease | ClinVar: VUS<br>gnomAD MAF: absent<br>Mayo PKDB: absent<br>Computational data: support pathogenicity<br>MyCode: absent<br>Less conservative changes to Ile, Lys, and Pro at the site have been considered pathogenic.<br>Residue is in PLAT domain of orthologs. | PM2<br>PM1<br>PM5<br>PP3 | Remains VUS. Inconclusive as there is limited evidence regarding pathogenicity of the variant; however, the available evidence makes this variant suspicious in nature. |
|  | NM_001009944.3:<br>c.3352G>T<br>p. Val1118Leu | CKD4, diagnosed in 60s<br>Cystic kidney disease<br>*TSC2* VUS<br>First degree relative with cystic kidney disease and a first degree relative with renal cell cancer. | ClinVar: absent<br>gnomAD MAF: low<br>Mayo PKDB: absent<br>Computational data: does not support pathogenicity<br>MyCode: absent<br>Alamut data noting mutational hot spot and/or critical domain | PM2<br>PM1<br>BP4 | Remains VUS. Inconclusive as there is limited evidence regarding pathogenicity of the variant. |
|  | NM_001009944.3:<br>c.11665G>T<br>p. Ala3889Ser | CKD2, diagnosed in 70s<br>Cystic kidney disease<br>No family history<br>*IFT140* VUS | ClinVar: absent<br>gnomAD MAF: low<br>Mayo PKDB: absent<br>Computational data: does not support pathogenicity<br>MyCode: 0/4 with ADPKD ICD codes<br>Alamut data noting mutational hot spot and/or critical domain | PM2<br>PM1<br>BP4 | Remains VUS. Inconclusive as there is limited evidence regarding pathogenicity of the variant. |
|  | NM_001009944.3:<br>c. 1910C>T<br>p. Ala637Val | Cystic kidney disease<br>FSGS<br>ESRD<br>Liver cysts<br>Dialysis, initiated in 40s<br>No family history | ClinVar: VUS<br>gnomAD MAF: absent<br>Mayo PKDB: LB<br>Computational data: does not support pathogenicity<br>MyCode: 0/1 with ADPKD ICD code; however, few left renal cysts noted on imaging.<br>Alamut data noting mutational hot spot and/or critical domain | PM2<br>PM1<br>BP4 | Remains VUS. Inconclusive as there is limited evidence regarding pathogenicity of the variant. |
|  | NM_001009944.3:<br>c.5830 G>A<br>p. Gly1944Arg | CKD3B, diagnosed in 70s<br>Cystic kidney disease<br>No family history | ClinVar: VUS<br>gnomAD MAF: low<br>Mayo PKDB: absent<br>Computational data: intermediate support<br>MyCode: 0/59 with ADPKD ICD code<br>Alamut data noting mutational hot spot and/or critical domain | PM2<br>PM1 | Remains VUS. Inconclusive as there is limited evidence regarding pathogenicity of the variant. |
| *PKD2* | NM_000297.4:<br>c.959G>T<br>p. Arg320Leu | ESRD, diagnosed in 70s<br>Cystic kidney disease<br>Multiple first and second-degree relatives with cystic kidney disease | ClinVar: absent<br>gnomAD MAF: absent<br>Mayo PKDB: absent<br>Computational data: supports pathogenicity<br>MyCode: 1/1 (100%) with ADPKD ICD code (OR 6219, 95% CI: 218, 177032; p<0.001)<br>Alamut data noting mutational hot spot and/or critical domain | PS4<br>PM2<br>PM1<br>PP3 | **Recommend upgrading to LP**, already done by GeneDx. |
|  | NM_000297.4:<br>c.901A>G<br>p. Thr301Ala | ESRD, diagnosed in 40s<br>Cystic kidney disease<br>*PKD1* variant<br>Multiple first and second-degree relatives with ADPKD | ClinVar: VUS<br>gnomAD MAF: low<br>Mayo PKDB: absent<br>Computational data: does not support pathogenicity<br>MyCode: 0/12 with ADPKD ICD code<br>Alamut data noting mutational hot spot and/or critical domain | PM1<br>BP4<br>BP5 | **Recommend downgrading** as MyCode data not supportive and computational prediction data not supportive. Proband's case is explained by *PKD1* pathogenic variant. |
| *IFT140* | NM_014714.4:<br>c.1126G>A<br>p. Glu376Lys | CKD2, diagnosed in 70s<br>Cystic kidney disease<br>Liver cysts<br>*PKD1* VUS<br>No family history | ClinVar: conflicting<br>gnomAD MAF: low<br>Mayo PKDB: absent<br>Computational data: does not support pathogenicity<br>MyCode: 45 participants in 34 families in total. None with ADPKD ICD-10 codes. | PM2<br>BP4 | Remains VUS. Inconclusive as there is limited evidence regarding pathogenicity of this variant. P/LP variants within *IFT140* results in a milder phenotype than PKD1/2 and have weaker penetrance. Difficult to conclude pathogenicity, particularly given the presence of *PKD1* VUS. Further, computational predictions are not supportive and there were no ICD-10 codes found in MyCode cohort. |
| *TSC1* | NM_000368.5:<br>c.475G>A<br>p. Gly159Ser | CKD1, diagnosed in 40s<br>Bilateral cystic kidney disease<br>Liver cysts<br>Pineal cyst<br>Nephrolithiasis<br>HTN<br>First degree relative with Budd Chiari syndrome<br>Second degree family history NOS kidney disease | ClinVar: absent<br>gnomAD MAF: low<br>LOVD: absent<br>Computational data: intermediate<br>MyCode: 2 participants in 2 families. 1 with bilateral renal cysts and 1 with renal angiomyolipoma. Participants were negative for other *TSC* features.<br>Alamut data noting mutational hot spot and/or critical domain | PM1<br>PM2 | Remains VUS. Inconclusive as there is limited evidence regarding pathogenicity of this variant, could have mild disease properties. Clinical case and MyCode participants did not meet clinical TSC diagnostic criteria. However, the clinical case did present with cystic kidneys and 1 MyCode participant has a personal history of angiomyolipoma. Further, computational predictions were intermediate. |
| *OFD1* | NM_003611.3:<br>c.44G>C<br>p. Ser15Thr | CKD3A, diagnosed in 70s<br>Cystic kidney disease<br>No family history | ClinVar: absent<br>gnomAD MAF: low<br>Computational data: Intermediate<br>MyCode: 30 participants (5 males, 25 females) in 22 families. None of the patients had ICD-10 codes for ADPKD. Nor did they have Joubert syndrome or Orofaciodigital syndrome diagnosis. | BS2<br>PM2 | Remains VUS. Inconclusive as there is limited evidence regarding pathogenicity of this variant. The computational data is intermediate and MyCode is not supportive. Additionally, none of the males in MyCode presented with OFD1-related concerns. Penetrance is suspected to be high; although, highly variable in expression with some affected females presenting with cystic kidneys. |

This table shows variants of uncertain significance (VUS) and any supportive evidence for or against pathogenicity for cystic kidney disease genes.
\*This column shows patients in the Geisinger MyCode 175k freeze with the VUS and case-control studies comparing ADPKD ICD diagnoses in VUS heterozygotes vs. non-carriers without rare variants in the corresponding genes for *PKD1*, *PKD2*, based on our prior work; <sup>32-34</sup> we excluded clinical cases who were in MyCode or were carrying an additional likely pathogenic PKD1/PKD2 variant [ClinVar, Mayo PKD Database, or PTV].
Abbreviations: LP (likely pathogenic), P (pathogenic), VUS (variant of uncertain significance), LB (likely benign), ESKD (end-stage kidney disease), AS (Alport syndrome), CKD (chronic kidney disease), HTN (hypertension), Mayo PKDB (Mayo PKD Database), LOVD (Leiden Open Variation Database), REVEL (Rare Exome Variant Ensemble Learner), ICD (International Classification of Diseases).

**Table 6.** Variants in Nephrolithiasis Genes (*SLC7A9, SLC34A3, OCRL*) Investigated.

| Nephrolithiasis |  |  |  |  |  |
| --- | --- | --- | --- | --- | --- |
| Gene | Variant | Clinical case | Variant details | ACMG criteria | Recommendations |
| *SLC7A9* | NM_014270.5:<br>c.177G>A<br>p. Thr59= | Nephrolithiasis<br>First degree family member with nephrolithiasis harboring the Phe140Ser variant. | ClinVar: conflicting<br>gnomAD MAF: absent<br>MyCode: absent | PM2 | Inconclusive as there is limited evidence regarding pathogenicity of this variant. |
|  | NM_014270.5:<br>c.419T>C<br>p. Phe140Ser |  | ClinVar: conflicting<br>gnomAD MAF: low<br>Computational data: supports pathogenicity<br>MyCode: 8/45 (17.8%) nephrolithiasis (OR 1.65, 95% CI: 0.76, 3.55; p=0.2) | PM1<br>PM2<br>PP3 | Inconclusive as there is limited evidence regarding pathogenicity of this variant. There were no homozygous controls within MyCode data. However, there was a trend toward higher risk with heterozygous. |
|  |  |  | Alamut data noting mutational hot spot and/or critical domain |  |  |
| SLC34A3 | NM_080877.2:<br>c.413C>T<br>p. Ser138Phe | Nephrolithiasis, diagnosed in 20s<br>No family history | ClinVar: conflicting<br>LOVD: absent<br>gnomAD MAF: low<br>Computational data: supports pathogenicity<br>MyCode: 128 participants compound heterozygous for p. Ser138Phe and p. Leu527del were found to have risk of nephrolithiasis ICD-10 code in 22.7% vs 11% in controls (p<0.001)<br>Functional testing: supports pathogenicity<br>Alamut data noting mutational hot spot and/or critical domain | PM1<br>PP2<br>PP3<br>PS3 | **Consider upgrading to LP** as computational predictions support pathogenicity as does functional testing. MyCode data is supportive of pathogenicity; although, there were no homozygous controls. |
|  | NM_080877.2:<br>c.1579_1581del<br>p. Leu527del |  | ClinVar: conflicting<br>LOVD: absent<br>gnomAD: low<br>MyCode: 128 participants compound heterozygous for p. Ser138Phe and p. Leu527del, among those with p.Leu527del without p.Ser138Phe, none had nephrolithiasis ICD-10 code vs 11% in controls (p=1.0) | PM2 | Inconclusive as there is limited evidence regarding pathogenicity of this variant. However, in the absence of p. Ser138Phe, there was no association with nephrolithiasis. |
|  | NM_080877.2:<br>c.304+2T>C<br>IVS4+2T>C (LP) | Nephrolithiasis<br>Hypercalciuria and phosphorus<br>Cystic kidney disease<br>Autism and learning disorder<br>Family history includes first degree relative with c.304+2T>C and nephrolithiasis | ClinVar: LP/P<br>gnomAD: present<br>Computational data: support pathogenicity<br>MyCode: nephrolithiasis noted in 16.9% vs. 11.2% in controls (OR 1.57, 95% CI: 1.01, 2.45; p=0.046) | PP3<br>PM1<br>PM3<br>PP4 | This variant is noted as pathogenic by the testing laboratory. |
|  | NM_080877.2:<br>c.561-8G>A<br>IVS6-8G>A<br>(VUS) |  | ClinVar: VUS<br>gnomAD: present<br>Computational data: intermediate<br>MyCode: 13.3% vs. 11.2% in controls (OR 1.39, 95% CI: 0.31, 6.28; p=0.7) | PP3<br>PM3 (likely) | Inconclusive: variant is present in population studies with intermediate computational analysis. MyCode data has limited evidence. |
| OCRL | NM_001318784.2:<br>c.2256+6T>C<br>Intronic | Bilateral nephrolithiasis thiazide resistant<br>Hypercalciuria<br>Hyperphosphatemia<br>High PTH<br>Sibling with nephrolithiasis and the same VUS as well as a sibling without nephrolithiasis and does not harbor the VUS | ClinVar: VUS<br>gnomAD MAF: low<br>Computational data: does not support pathogenicity<br>MyCode: 4 female participants in 4 families without ESRD ICD-10 code or nephrolithiasis ICD-10 code, and none had eGFR <60. Chart review for 2 revealed bilateral kidney cysts. 1 had a first degree relative with ESRD.<br>Segregation analysis: 1 proband and 1 segregate<br>Affects a nucleotide within consensus splice site | PM1<br>PM2<br>BP4 | Inconclusive as there is limited evidence regarding pathogenicity of this variant. While this variant is within a consensus splice site, computational algorithms do not suggest this variant affects RNA splicing. There were no male participants with this variant in the MyCode cohort, making it hard to assess the variant further. |

This table shows variants of uncertain significance (VUS) and any supportive evidence for or against pathogenicity for cystic kidney disease genes.
\*This column shows patients in the Geisinger MyCode 175k freeze with the VUS and case-control studies comparing ADPKD ICD diagnoses in VUS heterozygotes vs. non-carriers without rare variants in the corresponding genes for *PKD1*, *PKD2*, based on our prior work;<sup>32-34</sup> we excluded clinical cases who were in MyCode or were carrying an additional likely pathogenic PKD1/PKD2 variant [ClinVar, Mayo PKD Database, or PTV].
Abbreviations: LP (likely pathogenic), P (pathogenic), VUS (variant of uncertain significance), LB (likely benign), ESKD (end-stage kidney disease), AS (Alport syndrome), CKD (chronic kidney disease), HTN (hypertension), Mayo PKDB (Mayo PKD Database), LOVD (Leiden Open Variation Database), REVEL (Rare Exome Variant Ensemble Learner), ICD (International Classification of Diseases),

In other cases, data were suggestive but not conclusive per ACMG variant classification guidelines. For the *PKD1* c.7622 C>T (p.Pro2541Leu) variant, 0/65 had ADPKD ICD diagnoses in MyCode, it was absent from gnomAD but computational prediction data were intermediate. In an unresolved case, a patient with bilateral renal cysts, liver cyst, stage 1 CKD, and a family history of CKD in a second-degree relative was found to have a *TSC1* p.Gly159Ser VUS; REVEL (0.84), but not alpha missense (0.26) was supportive of pathogenicity. In our MyCode research cohort, there were 2 other individuals heterozygous for this variant with 1 having bilateral renal cysts and the other having a renal angiomyolipoma; neither had other tuberous sclerosis (TSC) features, though chart review was limited as none had been evaluated by a medical geneticist. Although the identified features in these 2 individuals can be seen in tuberous sclerosis, they are insufficient to meet TSC diagnostic criteria or ACMG variant classification criteria.^37^

Reclassification of the 10 VUS to LP has substantial implications for management. For example, one of the variants upgraded to LP, *HNF1B* c.907C>T (p.Arg303His; previously published^12^) had many downstream effects including kidney donor selection for 3 potential donors, family planning for 3 individuals, avoiding kidney biopsy in 1 individual, and extrarenal screening for low magnesium and diabetes in 3 individuals. Another individual who had nephrolithiasis beginning in his 20s and a family history of nephrolithiasis (second degree relative) was heterozygous for *SLC34A3* c.413C>T (p.Ser138Phe) and *SLC34A3* c.1579_1581del (p.Leu527del). Review of prior literature suggests that these 2 variants are part of a haplotype and inherited together.^38^ Our observations support the literature, as all *SLC34A3* c.413C>T (p.Ser138Phe) heterozygotes in MyCode carry the *SLC34A3* c.1579_1581del (p.Leu527del) variant. The risk of nephrolithiasis was higher in these individuals (22.7% vs. 11.2% in non-carriers; OR 2.23, 95% CI: 1.45, 3.44; p<0.001). The frequency of *SLC34A3* c.413C>T (p.Ser138Phe) in Gnomad is 0.00027, and alpha missense (0.83) and REVEL (0.67) scores are supportive of pathogenicity for this variant. None of the 7 individuals in MyCode heterozygous for *SLC34A3* c.1579_1581del (p.Leu527del) by itself had nephrolithiasis. These results suggest that *SLC34A3* c.413C>T (p.Ser138Phe) is pathogenic, though classification may fall outside of the traditional ACMG framework and could be considered as a likely risk allele.^39^

## Discussion

Our study demonstrates that the use of a large research biobank with exome sequence and medical records can provide evidence to support reclassification of pathogenicity in patients with rare VUS in kidney disease genes. This requires a careful multidisciplinary approach, collaborative work across institutions and expert groups. As similar large biobanks continue to expand, there is a valuable opportunity to use these data sources to resolve VUS and provide patients and families with important information to guide their care. Through these efforts we were able to provide evidence supporting reclassification 9 VUS to LP and 10 VUS to LB. We also found that in all *COL4A3/4/5* variants for which we had sufficient data, case-control analyses in MyCode were consistent with functional testing results by the Miner lab.

Collaborative approaches with kidney genetic experts and clinical genetic laboratories is critically important in resolving VUS. Ironically, on review of data and discussion with the clinical genetic testing laboratory, we noted that the *PKD1* variant c.7531G>C (p.Ala2511Pro) was likely double-counted by the testing genetic laboratory and in our publication and may need additional evidence for pathogenicity (**Table 5**).^32^

During the course of this study, data from MyCode and other biobanks were used to strengthen evidence for gene-disease association for heterozygous “carriers” of pathogenic variants in the genes *ALG8* and *SLC34A3.* ^16,18,19^. *ALG8* gene-disease validity classification was upgraded to “definitive” by the ClinGen Kidney Cystic and Ciliopathy Disorders panel based on MyCode data, and thus 3 patients with mild cystic kidney disease were reclassified as having *ALG8*-ADPKD, which has a much milder kidney and liver cystic phenotype with little risk of ESKD compared to ADPKD due to *PKD1/PKD2*.^16,40^ Our work illustrates how reclassification of VUS and recognition of disease risk related to heterozygous “carriers” provides opportunities for disease-specific care of patients.

We focused on the period beginning in 2020, when there was a clear increase in genetic testing in the nephrology department due to our efforts to improve recognition of genetic kidney disease and expedite genetic testing, driven by the growing awareness of the importance of rare genetic causes of kidney disease.^1,2,5,7,41,42^ The KDIGO held a Controversies Conference on Genetics in March 2021 and emphasized the importance of practitioners to adopt a “think genetic” approach, encouraging them to obtain family history, onset of CKD, evaluating for extrarenal symptoms, and considering genetic testing.^1^ ^42^ The median time from kidney disease diagnosis to genetic testing in our cohort was 8 years. With increasing availability and uptake, genetic testing may help reduce the diagnostic odyssey for individuals with CKD.

Timely genetic testing in kidney disease is crucial, as it can have important clinical implications for patient care as shown in our study and others.^2,7,43,44^ Benefits include more accurate diagnosis, personalized treatment, screening for extrarenal conditions, family planning, and identification of appropriate living-related kidney donors. In an Australian study with 204 nephrology patients, 39% had a change in clinical diagnosis, 59% had a change in management, and 20% had a change in treatment plan after exome sequencing.^44^ In a 2023 US study, CKD-focused genetic testing was conducted on 1623 CKD patients, of which 20.8% had positive genetic findings. Of those patients who had positive results, 48.8% had a change in diagnosis, 90.7% had a change in management, and 32.9% had a change in treatment plan. Certain pre-test indications had higher diagnostic yield than others though even indications with lower diagnostic yield may benefit given important impacts on management. For example, Anderegg et al. found that the diagnostic yield for monogenic disease in an unselected nephrolithiasis cohort was 2.9% (23/787) though an additional 64 (8.1%) were monoallelic for likely pathogenic variants predisposing to nephrolithiasis.^45^ In our study, 2/24 (8.3%) of patients tested for nephrolithiasis or nephrocalcinosis had a monogenic cause with an additional 1 patient having a VUS where we had suggestive evidence to upgrade (*SLC34A3* c.413C>T (p.Ser138Phe) and 2 more patients with suspicious VUS that we were unable to resolve (**Table 6**). As previously observed, there is evidence to support a 2-3-fold higher risk associated with *SLC34A3* variants in the heterozygous state, and there is the potential for personalization by increasing phosphate intake by supplementation to decrease the stimulus for calcitriol production and hyperabsorption of calcium.^18,46,47^

While some cases did not result in changes to management, we highlight the importance of ending the patient’s “diagnostic odyssey” and the significance of providing the patient with an explanation for their CKD.^42,48^ Identifying the cause for CKD provide patients with opportunities, such as the potential for enrollment into clinical trials. Genotype-based precision clinical trials are underway for various genetic kidney diseases, and patients with known genetic diagnoses could become eligible for existing or future trials. Additionally, the role of genetic testing in living kidney donation is particularly important as we found that 2 kidney donors who later developed CKD5/ESKD had monogenic causes, including one with ADTKD-UMOD^49^ and the other with autosomal dominant Alport syndrome.

Recommendations for living kidney donors have been made by kidney transplant experts as well as the aforementioned NKF multidisciplinary working group.^42,43^ Foremost, patients being considered for transplant with CKD of unknown cause or suggestive features (e.g. extrarenal findings, syndromic presentation) should receive genetic testing to help identify potentially at-risk family members and avoid transplantation of a kidney that is susceptible to disease. However, there is controversy about whether high-risk genotype with intermediate (approximately 2-fold) risk of CKD such as *APOL1* and intermediate-effect genetic conditions such as autosomal dominant Alport syndrome should be absolute contraindications for transplantation. The transplant review suggests using shared decision-making about the decision to proceed with transplant.^43^

There were several limitations in our study. Most notably, this was restricted to a single center. Future efforts should be made to replicate this approach across other centers, ideally leveraging larger biobanks with high quality phenotyping data. Additionally, we were unable to ascertain the specific rationale for the type of genetic testing being ordered. This decision was likely dependent on provider preference, insurance coverage, and/or patient preference. In particular, patients’ desire to pursue comprehensive testing may reflect on their understanding of monogenic forms of kidney disease as well as their comfort level with receiving uncertain results given the option to forego VUS reporting per laboratory policy. Genetic presentations of kidney disorders are often highly heterogenous, and further research is needed to determine optimal testing strategies that maximize diagnostic yield while minimizing overburdensome VUS reporting. On the other hand, integrating broad-based panel testing with rigorous, multidisciplinary evaluation could offer a powerful approach to resolving many of these cases.

In conclusion, we demonstrate the utility of a multidisciplinary approach that leverages biobank data integrating exome sequencing with electronic health records, along with functional testing and collaboration with genetic testing labs, to enhance reclassification of clinically significant VUS.

## Supporting information

MyCode Supplement

## Data Availability

All data produced in this study will be made available by emailing the corresponding author.

## Acknowledgements

We would like to acknowledge Joshua Begin for assistance with trimerization assays. We are thankful to the Geisinger Nephrology and Genetics Departments in providing assistance in identifying patients who underwent genetic testing. We are grateful to the many patients who contributed to the MyCode Community Health Initiative by providing genomic and electronic health information. We thank the Regeneron Genetics Center for providing funding for patient enrollment and exome sequencing for the DiscovEHR study, and we would like to acknowledge the Geisinger-Regeneron DiscovEHR Collaboration for the genotypic and phenotypic data. This work was supported by NIH NHGRI R01HG011799. VUS analysis in the Washington University Kidney O’Brien Center for Chronic Kidney Disease Research Variant Validation Core was supported by NIH grant U54DK137332 to JHM.

