## Supplementary material for "Leveraging genomic biobanks to enhance genetic testing outcomes for kidney disease": MyCode Supplement

| Transcript | Gene | Cdot | Pdot | N (number of families) |
|---|---|---|---|---|
| | Controls* | | | |
| NM_001009944.3 | PKD1 | 7479C>G | Phe2493Leu | 15 (13) |
| NM_001009944.3 | PKD1 | 12862A>G | Ser4288Gly | 65 (54) |
| NM_001009944.3 | PKD1 | 8293C>T | Arg2765Cys | 2566 (2344) |
| NM_001009944.3 | PKD1 | 3568C>T | Arg1190Cys | 12 (9) |
| NM_001009944.3 | PKD1 | 7622 C>T | Pro2541Leu | 65 (56) |
| NM_001009944.3 | PKD1 | 11665G>T | Ala3889Ser | 5 (5) |
| NM_001009944.3 | PKD1 | 1910C>T | Ala637Val | 1 (1) |
| NM_001009944.3 | PKD1 | 5830 G>A | Gly1944Arg | 59 (50) |
| NM_000297.4 | PKD2 | 959G>T | Arg320Leu | 1 (1) |
| NM_000297.4 | PKD2 | 901A>G | Thr301Ala | 12 (9) |

*Controls defined as those without rare variants in PKD1 or PKD2 at allele frequency <0.01

Firth logistic regression analyses adjusted for age, sex, race, ethnicity, initial year; excludin

ADPKD diagnosis defined as having 1 or more of the following *ICD-9* or *ICD-10* codes in electro

| ADPKD ICD code<br>n/N (%) | OR (95% CI:); p val | Any kidney or liver cystic ICD code<br>n/N (%) | OR (95% CI:); p value |
|---|---|---|---|
| 13/19265 (0.07%) | Ref | 578/19265 (3.0%) | Ref |
| 0/15 | N/A | 0/15 | N/A |
| 0/64 | N/A | 1/64 (1.6%) | 0.74 (0.15, 3.76); p=0.7 |
| 2/2533 (0.08%) | 1.45 (0.38, 5.59); p= | 72/2533 (2.8%) | 0.97 (0.75, 1.24); p=0.8 |
| 0/11 | N/A | 0/11 | NA |
| 0/65 | N/A | 4/65 (6.2%) | 2.41 (0.91, 6.38); p=0.08 |
| 0/5 | N/A | 0/5 | N/A |
| 0/1 | N/A | 0/1 | N/A |
| 0/59 | N/A | 2/59 (3.4%) | 1.45 (0.40, 5.21); p=0.6 |
| 1/1 (100%) | 6219 (218, 177032) | 1/1 (100%) | 84.51 (3.43, 2079.53) |
| 0/12 | N/A | 0/12 | N/A |

.

ıg clinical cases who were in MyCode as well those harboring other PKD1/PKD2 P/LP variants in ClinVar, Ma
nic health record: Q61.2, Q61.3, 753.12, or 753.13.

ıyo PKD Database, or protein-truncating variants
