## Supplemental Material for "Leveraging genomic biobanks to enhance genetic testing outcomes for kidney disease"

**Supplemental Materials.**

**Supplemental Methods**

**Supplemental Table 1. Power Calculations for Case-control Association Analyses**

**Supplemental Table 2. Genetic Testing Outcomes**

**Supplemental Table 3. Examples of Implications of Genetic Testing by Diagnostic Indication among Positive-tested Patients**

**Supplemental Table 4. Variants in Other Genes (*WT1, MYH9)* Investigated**

**Supplemental Figure. In vitro splicing analysis of *COL4A5* c.937-10T>A**

**Supplemental Table 5. Assessments of VUS according to Alamut version 1.13 (prior to our analysis)**

### Supplemental Methods

#### Splicing reporter minigene assay

The splicing reporter minigenes for human COL4A3 WT and two variants (c.987+2T>C and c.987G>A) were generated by synthesizing exon 17 sequence plus 158 and 168 bp of flanking intronic sequences and inserting into the pCAS2.1 splicing vector via *BamHI* and *MluI* restriction sites (Gaildrat P, 2010) (GenScript Corp., Piscataway, NJ). The splicing assay was performed as described previously (Malone A, 2016) with some modifications. HeLa cells were seeded on a 12-well plate ($5x10^4$ cells/well) in DMEM (Thermo Fisher Scientific, Waltham, MA) supplemented with 10% FBS and penicillin-streptomycin and cultured overnight at 37°C with 5% $CO_2$ to about 60% confluency. Cells were transfected with the empty vector, WT vector, variant vector, or WT plus variant vectors using Lipofectamine 2000 reagent (Invitrogen, Carlsbad, CA) with 1 µg DNA total in each well. At 20 h after transfection RNA was extracted with an RNeasy Micro kit (Qiagen, Hilder, Germany). One µg of each RNA sample was subjected to cDNA synthesis with PrimeScript RT Master Mix (Takara Bio, Mountain View, CA) followed by 35 cycles of PCR amplification with an annealing temperature at 55C using PrimeStar Max DNA polymerase (Takara Bio). The primers used for PCR were pCAS-KO1-Forward primer (5'-TGACGTCGCCGCCCATCAC-3') and pCAS-2-Reverse primer (5'-ATTGGTTGTTGAGTTGGTTGTC-3'). The amplicons were then resolved by electrophoresis on a 2% agarose gel to determine the splicing patterns. Bands of interest were purified from the gel using a Wizard SV Gel and PCR Clean-Up kit (Promega, Madison, WI) and subjected to Sanger sequencing (GENEWIZ/Azenta Life Sciences, South Plainfield, NJ) with the pCAS-KO1-Forward primer.

#### Trimerization reporter NanoLuciferase assay

The split NanoLuciferase-based trimerization assay was performed as previously described using the human COL4A4 plasmid, the human COL4A3 and COL4A5 plasmids with N-terminal split NanoLuciferase tags, and the firefly luciferase plasmid for normalization) (Omachi et al. Cell Chem Biol. PMID: 29526710). The COL4A3 G1219C (c.3655G>T) (Pescucci et al. Kidney Int. 2004; PMID: 15086897), COL4A4 G960R (c.2878G>C) (Kohler et al. Kidney360 2022; PMID 36514391), and COL4A5 G521D (c.1562G>A) (Omachi et al. Cell Chem Biol. PMID: 29526710) variants were used as pathogenic controls. Each variant tested was introduced into its respective COL4A cDNA plasmid by GenScript (Piscataway, NJ). Dual luciferase assays were performed in transfected HeLa cells as described (Yu S et al.

Kidney Int. 2024; PMID 38782199) except that each transfection was done in a single well of 6-well plates and replated in 6 replicates on 96-well plates. VUS were identified as pathogenic if less than 75% of WT luminescence in intracellular and/or extracellular assays was observed.

**Supplemental Table 1. Power Calculations for Case-control Associations**

| | Minimal sample size of VUS carriers | |
|---|---|---|
| | 80% power | 90% power |
| *PKD1/PKD*2 – ADPKD ICD* | | |
| 90% prevalence | 4 | 4 |
| 80% prevalence | 5 | 6 |
| 70% prevalence | 6 | 8 |
| *COL4A3/4/5* – dipstick hematuria** | | |
| 3-fold higher prevalence (63%) | 21 | 27 |
| 2.5-fold higher prevalence (52.5%) | 36 | 48 |
| 2-fold higher prevalence(42%) | 76 | 101 |
| *SLC34A3* – nephrolithiasis ICD*** | | |
| 2.5-fold higher prevalence (27.5%) | 89 | 118 |
| 2-fold higher prevalence (22%) | 178 | 238 |
| 1.5-fold higher prevalence (16.5%) | 615 | 822 |

Assumes 0.14% prevalence ADPKD, 21% prevalence dipstick hematuria in controls.

Estimates for PKD1/PKD2 and COL4A3/4/5 based on MyCode 175k freeze data: PMID **36573973**, **37849993, 39625784**

*ADPKD diagnosis defined as having 1 or more of the following *ICD-9* or *ICD-10* codes in electronic health record: Q61.2, Q61.3, 753.12, or 753.13.

**Dipstick hematuria defined as having trace blood or greater (i.e., trace, 1+, 2+, 3+) on at least half of all urinalyses, excluding urinalyses positive for leukocyte esterase or nitrites

***Nephrolithiasis diagnosed by phecode GU_585, which includes ICD-10 codes (N20, N20.0, N20.1, N20.2, N20.9, N21, N21.0, N21.1, N21.8, N21.9, N22, V13.01) and ICD-9 codes (274.11, 592, 592.1, 592.9, 594, 594.1, 594.2, 594.8, 594.9)

**Supplemental Table 2. Genetic testing outcomes**

| Genetic testing outcomes for patient | |
|---|---|
| Clinical domain | Explanation and Example |
| Change in clinical management:<br>• Avoidance of additional testing<br>• Avoidance of unnecessary therapy<br>• Personalized treatment plan<br>• Management of extrarenal manifestations | Results directly changed patient's management:<br>• Avoidance of kidney biopsy<br>• Avoidance of steroids for FSGS due to genetic cause<br>• Informed tolvaptan use, tailored nephrolithiasis management<br>• Screening for diabetes and hypomagnesemia (*HNF1B*), hearing, ocular abnormalities (*COL4A3/4/5*) |
| Refine diagnosis:<br>• Confirmed diagnosis<br>• Reclassified diagnosis<br>• Prognostic clarification | Facilitates or confirms genetic diagnosis<br>• Facilitates or confirms genetic diagnosis<br>• Reclassifies disease from either incorrect diagnosis or specific diagnosis to genetic diagnosis (e.g. CKDu to Alport Syndrome; atypical cystic kidney disease to ADPKD due to *ALG8, GANAB*, etc.)<br>• Genotype information for PROPKD score, more specific genetic diagnosis for FSGS |
| Genetic testing outcomes for family members | |
| Clinical domain | Explanation of domain |
| At risk relatives identified:<br>• Identified possible affected relatives<br>• Better awareness of reproductive risk | Family sharing to inform relatives of their risk<br>• Cascade testing and/or clinical screening<br>• Carrier testing to provide accurate reproductive risk |

### Supplemental Table 3. Examples of Implications of Genetic Testing by Diagnostic Indication among Positive-tested Patients

| **Diagnostic Indication** | **Clinical management changes for patient N (%)** | **Facilitate, refine, or reclassify diagnosis N (%)** | **Familial implications N (%)** | **Examples** |
|---|---|---|---|---|
| Overall | 77/228 (34%) | 73/78 (93.6%) | 139/228 (61%) | See below |
| Cystic kidney disease | 32/69 (54%) | 44/69 (64%) | 42/69 (61%) | *PKD1, PKD2, HNF1B, OFD1, ALG8, TSC2, COL4A1, COL4A3, COL4A4, SLC34A3, GANAB, SEC63*: Confirm diagnosis in atypical or early-onset cases; inform prognosis and use of tolvaptan and other appropriate treatments; screen for extrarenal manifestations; identify potential genetic risk for related kidney donors |
| CKDu | 12/40 (30%) | 15/40 (38%) | 23/40 (58%) | *COL4A4, COL4A5, CD2AP, UMOD, PAX2, MUC1, LMNA, EYA1,* 17q12 syndrome *(HNF1B), APOL1* high-risk allele: inform appropriate treatment depending on gene identified; screen for extrarenal manifestations, minimize unnecessary hematuria workup (*COL4A4, COL4A5*); avoid unnecessary immunosuppression (e.g. in FSGS); identify potential genetic risk for related kidney donors |
| Nephrolithiasis/ nephrocalcinosis | 3/24 (13%) | 3/24 (13%) | 8/24 (33%) | *SLC3A1* (biallelic), *CYP24A1* (biallelic), *SLC34A3* (heterozygous suspicious VUS)*;* personalized therapy based on gene; avoid excessive vitamin D and calcium supplements for conditions predisposing to hypercalciuria |
| FSGS or SRNS | 5/21 (24%) | 5/21 (24%) | 13/21 (62%) | *APOL1* high-risk allele, *COL4A3*, *INF2*, *NPHS2*: avoidance of unnecessary immunosuppression; minimize unnecessary hematuria workup (*COL4A3*); identify potential genetic risk for related kidney donors |
| Donor | 2/12 (17%) | 1/12 (8%) | 8/12 (67%) | *HNF1B, CFI*: inform shared decision-making around kidney donation eligibility; screen for extrarenal manifestations (e.g. hypomagnesemia, diabetes for *HNF1B*) |
| Suspected Alport syndrome | 7/11 (63.6% | 8/11 (73%) | 8/11 (73%) | *COL4A3 and COL4A4 ADAS, X-linked COL4A5*: Early initiation of RAAS inhibitors to delay risk of kidney failure; extrarenal screening for hearing loss and ocular abnormalities; minimize unnecessary hematuria workup, avoid unnecessary immunosuppression (e.g. in FSGS); identify potential genetic risk for related kidney donors |
| Hypertension/ nephrosclerosis | 2/11 (18%) | 3/11 (27%) | 9/11 (82%) | *APOL1 high-risk allele, TTR*: consider personalized treatment for amyloidosis; avoid unnecessary immunosuppression (e.g. in FSGS); evaluate for extrarenal conditions; identify potential genetic risk for related kidney donors |
| Glomerular | 3/10 (30%) | 3/10 (30%) | 1/10 (10%) | *COL4A3, COL4A4, COL4A5, PKD1*: Early initiation of RAAS inhibitors to delay risk of kidney failure; screening for extrarenal manifestations; avoid unnecessary immunosuppression; identify potential genetic risk for related kidney donors |
| Tubulointerstitial | 4/7 (57%) | 4/7 (57%) | 7/7 (100%) | *UMOD, COL4A3:* Early initiation of RAAS inhibitors to delay risk of kidney failure; extrarenal screening for hearing loss and ocular abnormalities; minimize unnecessary hematuria workup, avoid unnecessary immunosuppression (e.g. in FSGS) |
| ESKD w/DM | 0/7 | 0/7 | 4/7 (57%) | N/A |
| CAKUT | 0/6 | 0/6 | 3/6 (50%) | N/A |
| Hematuria | 1/5 (20%) | 1/5 (20%) | 2/5 (40%) | *COL4A5 suspicious VUS*: Early initiation of RAAS inhibitors to delay risk of kidney failure; screening for extrarenal manifestations; avoid unnecessary immunosuppression; identify potential genetic risk for related kidney donors |
| Atypical HUS | 1/3 (33%) | 1/3 (33%) | 2/3 (67%) | *CFH*: atypical HUS: anticipate using eculizumab during peritransplant period |
| Family Hx | 0/2 | 0/2 | 0/2 | N/A |

**Supplemental Table 4. Variants in Other Genes (*WT1, MYH9)* Investigated**

| Syndromic | | | | | |
|---|---|---|---|---|---|
| Gene | Variant | Clinical case | Variant details | ACMG criteria | Recommendations |
| *WT1* | NM_024426.6:<br>c.779C>T<br>p. Ser260Leu | • CAKUT<br>• ESRD<br>• Unilateral cystic kidney disease<br>• Hypospadias<br>• Child with hypospadias, variant status is unclear<br>• Multiple first-degree family members with NOS kidney disease | • ClinVar: VUS<br>• gnomAD MAF: Low<br>• Computational data: does not support pathogenicity<br>• MyCode: 21 participants in 17 families. One participant with ESRD diagnosed at 30-34 years of age due to vesicoureteral reflux (p=0.3). 16% with eGFR <60 vs 20% with eGFR <60 (p=1.0). On chart review, no Wilms' tumor or obvious WT1-related genitourinary abnormalities<br>• Alamut data noting mutational hot spot and/or critical domain | • PM1<br>• PM2<br>• BP4 | Inconclusive as there is limited evidence regarding pathogenicity of this variant. Computation predictions are not supportive of pathogenicity and 1 MyCode participant with ESRD without other obvious features per chart review. |
| *MYH9* | NM_002473.6:<br>c.2958G>C<br>p. Gln986His | • FSGS, diagnosis at a "young age"<br>• Nephrotic syndrome<br>• Dialysis<br>• 3rd degree relative with Alport syndrome | • ClinVar: conflicting<br>• gnomAD MAF: low<br>• LOVD: absent<br>• Computational data: supports pathogenicity<br>• MyCode: absent<br>• Alamut data noting mutational hot spot and/or critical domain | • PM1<br>• PM2<br>• BP4<br>• | Inconclusive as there is limited evidence regarding pathogenicity of this variant. |
| | NM_002473.6:<br>c.4024C>T<br>p. Arg1342Trp | • ESRD, diagnosed in 40s<br>• Proteinuria<br>• Dialysis<br>• s/p KT<br>• *APOL1* G1/G1 | • ClinVar: conflicting<br>• gnomAD MAF: low<br>• Computational data: does not support pathogenicity<br>• MyCode: 3 participants in 3 families. 1 with eGFR <60 (p=0.4), 2 with proteinuria (p=0.1)<br>• Alamut data noting mutational hot spot and/or critical domain | • PM1<br>• PM2<br>• BP4<br>• | Inconclusive as there is limited evidence regarding pathogenicity of this variant. |
| | NM_002473.6:<br>c.60G>T<br>p. Pro20= | • FSGS, diagnosed at young age<br>• ESRD, diagnosed in 20s<br>• IgM Nephropathy<br>• MCD<br>• *COL4A3* VUS | • ClinVar: absent<br>• gnomAD MAF: low<br>• LOVD: absent<br>• Computational data: NA<br>• MyCode: 7 participants in 7 families, none with ESRD ICD-10 code or FSGS ICD-10 code. None of the patients had eGFR <60. | • PM2 | Inconclusive as there is limited evidence regarding pathogenicity of this variant. Limited data including synonymous change, and MyCode data is not supportive, though small numbers. |

**Supplemental Figure. In vitro splicing analysis of *COL4A5* c.937-10T>A**

in vitro splicing analysis provided clear evidence that this variant induces aberrant splicing. Specifically, we observed:

- ✓ a splicing pattern with an additional 8 nucleotides derived from part of intron 16 (pattern ②),
- ✓ a splicing pattern with an additional 92 nucleotides from intron 16 (pattern ③), and
- ✓ an unidentified sequence (pattern ④).

Based on these findings, we concluded that this variant is a pathogenic variant causing X-linked Alport syndrome.

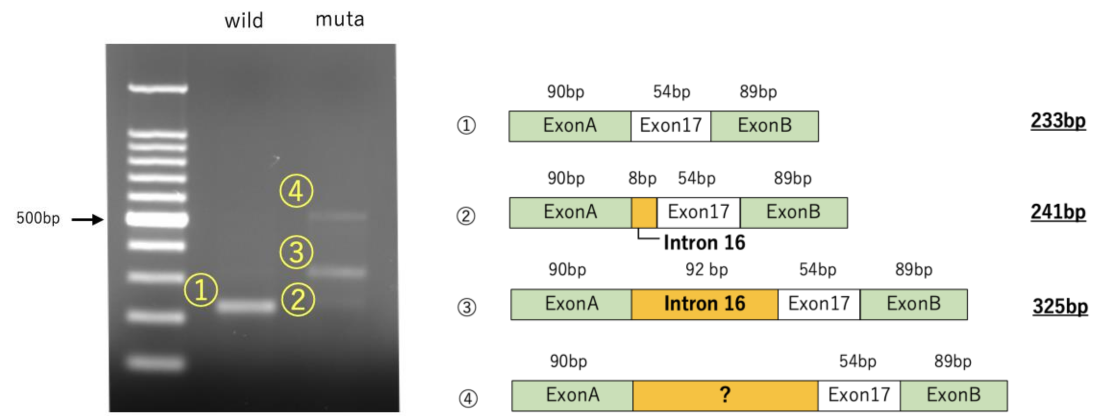


Wild Type

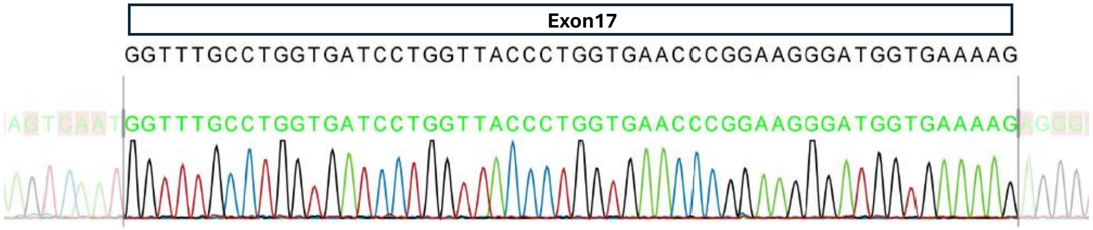


c.937-10T>A

②

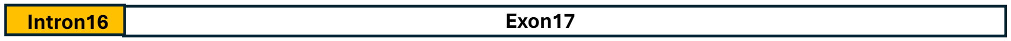

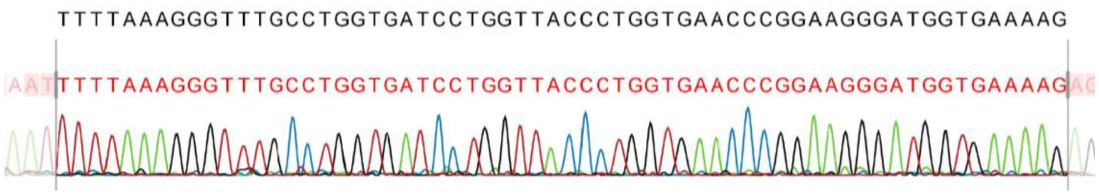
TTTTAAAGGGTTTGCCTGGTGATCCTGGTTACCCTGGTGAACCCGGAAGGGATGGTGAAAAG
TTTTAAAGGGTTTGCCTGGTGATCCTGGTTACCCTGGTGAACCCGGAAGGGATGGTGAAAAG

③

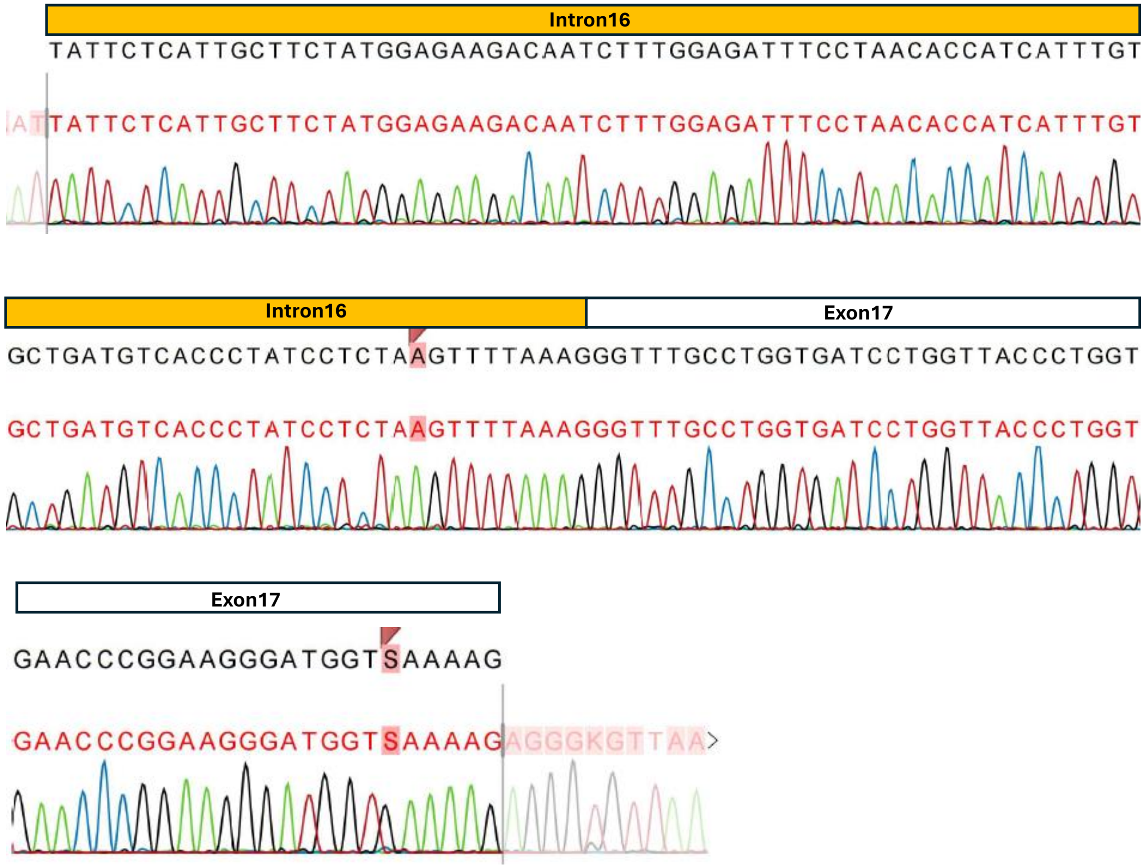
Intron16
TATTCTCATTGCTTCTATGGAGAAGACAATCTTTGGAGATTTCCTAACACCATCATTTGT
TATTCTCATTGCTTCTATGGAGAAGACAATCTTTGGAGATTTCCTAACACCATCATTTGT
Intron16
Exon17
GCTGATGTCACCCTATCCTCTAAGTTTTAAAGGGTTTGCCTGGTGATCCTGGTTACCCTGGT
GCTGATGTCACCCTATCCTCTAAGTTTTAAAGGGTTTGCCTGGTGATCCTGGTTACCCTGGT
Exon17
GAACCCGGAAGGGATGGTSAAAAG
GAACCCGGAAGGGATGGTSAAAAG

**Supplemental Table 5. Assessments of VUS according to Alamut version 1.13 (prior to our analysis)**

| Gene-disease association (Mode of inheritance) | Variant, relevant publications | Other Relevant Databases, Alamut score v.1.13 |
|---|---|---|
| *PKD1* – ADPKD (AD) | NM_001009944.3:c.7479C>G (p.Phe2493Leu) | Mayo PKD – not listed;<br>VUS (PM1_Moderate,PM2_Moderate ,PP3_Supporting) score = 5 |
| | NM_001009944.3:c.6101G>T (p.Gly2034Val)<br>Previously published in PMID **36573973** | Mayo PKD – not listed. Seen in 1 individual per communication via email. Well conserved site in orthologs and the PKD repeats; would consider LP<br>VUS (PM1_Moderate,PM2_Moderate, PP3_Supporting) score = 5 |
| | NM_001009944.3:c.7531G>C (p.Ala2511Pro)<br>Previously published in PMID **36573973** | Mayo PKD – VUS<br>VUS (PM1_Moderate, PM2_Moderate, BP4_Supporting) score = 3 |
| | NM_001009944.3:c.12862A>G (p.Ser4288Gly) | Mayo PKD – likely benign;<br>VUS (PM2_Moderate,BP4_Supporting) score = 1 |
| | NM_001009944.3:c.8293C>T (p.Arg2765Cys)<br>Many such as PMID 19165178 | Mayo PKD – VUS ; VUS (PM1_Moderate,PP3_Supporting) score = 3 |
| *PKD1* – ADPKD (AD) | NM_001009944.3:c.7622 C>T (p.Pro2541Leu) | Mayo PKDB – not listed;<br>VUS (PM1_Moderate,PM2_Moderate,PP3_Supporting) score =5 |
| *PKD1* – ADPKD (AD) | NM_001009944.3:c.1543G>A p.(Gly515Arg);<br>PMID 23985799 | Mayo PKDB – Likely pathogenic; VUS (PM1_Moderate,PM2_Moderate, PP3_Supporting) score = 5 |
| *PKD1* – ADPKD (AD) | NM_001009944.3:c.3568C>T p.(Arg1190Cys) | Mayo PKDB – not listed; VUS (PM1_Moderate,PM2_Moderate, BP4_Supporting) score = 3 |
| *PKD2* – ADPKD (AD) | *PKD2* NM_000297.4:c.959G>T (p.Arg320Leu) | Mayo PKD – not listed but 1 patient with Arg320Gln; VUS (PM1_Moderate, PM2_Moderate, PP3_Supporting) score = 5 |
| | *PKD2* NM_000297.4:c.901A>G (p.Thr301Ala) | Mayo PKD – not listed; VUS (PM1_Moderate,PM2_Moderate,BP4_Supporting) score = 3 |
| *COL4A4* – Alport Syndrome (AD/AR) | *COL4A4* NM_000092.5:c.2629C>T (p.Arg877Trp)<br>PMID **35485766** | LOVD – Not classified; VUS (PM1_Moderate,PM2_Moderate,BP4_Supporting) score = 3 |
| | *COL4A4* NM_000092.5:c.1055C>T (p.Pro352Leu) | LOVD – Conflicting (1 VUS, 1 Likely benign); VUS (PM1_Moderate,PM2_Moderate,PP3_Supporting,BP4_Supporting ) score = 4 |
| *COL4A3* – Alport Syndrome (AD/AR) | *COL4A3 c.520G>A (p.Gly174Arg)*<br>PMID 35419377 | LOVD – likely pathogenic; VUS (PM1_Moderate,PM2_Moderate,PP3_Supporting) score = 5 |
| *COL4A3* – Alport Syndrome (AD/AR) | *COL4A3* NM_000091.5:c.3644G>A (p.Arg1215Gln); | LOVD – Conflicting (1 LP, 2 likely benign, 1 not classified) VUS (PM1_Moderate,PM2_Moderate,BP4_Supporting) score = 3 |

| | | |
|---|---|---|
| | Several publications such as PMID **15954103** | |
| *COL4A3* – Alport Syndrome (AD/AR) | *COL4A3* NM_000091.4:c.987G>A (p.Lys329=) | LOVD – not listed; VUS (PM2_Moderate,PP5_Supporting, BP4_Supporting) score = 2 |
| *COL4A5* – Alport Syndrome (XL) | *COL4A5* NM_000495.5:c.4700G>T (p.Cys1567Phe) | LOVD – not listed; VUS (PM1_Moderate,PM2_Moderate,PP3_Supporting) score = 5 |
| | *COL4A5* NM_000495.4:c.937-10T>A (intronic) | LOVD – not listed; VUS (PM2_Moderate), score = 2 |
| *HNF1B – ADTKD-HNF1B* (AD) | *HNF1B* NM_000458.4:*c.908G>A (p.Arg303His)*<br>*PMID 36090499* | Likely pathogenic;<br>VUS (PM1_Moderate,PM2_Moderate,PP3_Supporting) score = 5 |
| *SLC34A3* - – *hypophosphatemic rickets with hypercalciuria (AR)* | *SLC34A3,* NM_001177316.2 *:* c.413C>T (p.Ser138Phe)<br>PMID 22159077, 26399350 | Not listed in LOVD; VUS (PM1_Moderate,PM2_Moderate,PP3_Supporting) score = 5 |
| | *SLC34A3*<br>NM_001177316.2*:*c.1579_1581del (p.Leu527del)<br>PMID 16358214, 24700880 | VUS (PM2_Moderate) score = 2 |
| | c.304+2T>C IVS4+2T>C (LP) | LOVD – absent<br><br>LP (PVS1, PM2, PP5) |
| | c.561-8G>A IVS6-8G>A (VUS) | LOVD – absent<br><br>VUS (PM2) |
